# The cellular hierarchy of acute myeloid leukemia informs personalized treatment

**DOI:** 10.1101/2024.07.24.24310768

**Authors:** Yannik Severin, Yasmin Festl, Tobias M. Benoit, Rebekka Wegmann, Benjamin D. Hale, Michael Roiss, Anne-Kathrin Kienzler, Thomas Pabst, Michael Scharl, Shinichi Sunagawa, Markus G. Manz, Antonia M.S. Müller, Berend Snijder

## Abstract

Acute myeloid leukemia (AML) is characterized by malignant myeloid precursors that span a cellular hierarchy from dedifferentiated leukemic stem cells to mature blasts. While the diagnostic and prognostic importance of AML blast maturation is increasingly recognized, personalized therapies are currently not tailored to a patients individual makeup of this cellular hierarchy. In this study, we use multiplexed image-based *ex vivo* drug screening (pharmacoscopy) to systematically quantify the drug sensitivity across the cellular hierarchy of AML patients. We analyzed 174 prospective and longitudinal patient samples from 44 newly diagnosed AML patients, which indicated that differences in the AML hierarchy significantly identified poor responses to first-line therapy, outperforming European LeukemiaNet (ELN) criteria. Critically, drug response profiling across the AML hierarchy of each patient improved the accuracy of predicting patient response to first-line therapy (AUC 0.91), and revealed alternative individualized treatment options targeting the complete AML hierarchy of non-responding patients. We confirmed these findings in an independent cohort of 26 relapsed/refractory AML patients, for whom pan-hierarchy response profiling improved response predictions *post hoc*. Overall, our results quantify the clinical importance of therapeutically targeting the complete cellular hierarchy of newly diagnosed AML, and identify multiplexed image-based *ex vivo* drug screening to enable quantification and targeting of the AML maturation hierarchy for improved personalized treatment.

## Introduction

Acute myeloid leukemia (AML) is a heterogeneous hematologic malignancy, in which complex combinations of chromosomal aberrations and somatic mutations lead to the rapid clonal expansion of immature myeloid cells ^1–3^. AML is characterized by the presence of a cellular hierarchy comprising multiple malignant cellular phenotypes reflecting varying levels of differentiation: Leukemias comprised of more mature blasts correlate with better prognosis and responses to treatment ^4–7^, whereas leukemias with minimal maturation are afflicted with lower overall survival ^4–11^.

AML treatment remains challenging due to drug resistance and relapse, which are often caused by the persistence of the most immature AML population, the leukemic stem cells (LSCs). Traditional AML treatment regimes struggle to eliminate LSCs and an incomplete eradication of these highly treatment-resistant cells can contribute to increased disease complexity, further complicating subsequent therapeutic efforts ^12,13^. Despite the inherent heterogeneity of AML, most patients receive similar standard-of-care treatments, since only a small proportion of AML patients have targetable mutations that can be treated with small molecules ^1^. This contributes to inconsistent treatment outcomes and varying overall survival rates ^14^. This underscores the need for personalized treatment approaches that target both LSCs and the leukemic bulk, while taking into account the distinct genetic and phenotypic variations present in each patient’s disease. By tailoring therapy to these characteristics, clinicians may be able to achieve long-term remission, improve treatment success and ultimately enhance patient survival^15^.

In recent years, there has been a growing interest in identifying and targeting molecular vulnerabilities unique to individual AML patients. Genomic testing uses recurrent genetic patterns to stratify AML patients into subgroups characterized by distinct clinical outcomes ^16^. However, genomic testing as performed in clinical routine often falls short, in part due to its inability to quantify the intratumoral heterogeneity related to blast differentiation ^17^. *Ex vivo* drug response profiling, also called functional precision medicine, complements such genomic associations by providing a direct functional characterization of the patient’s disease. Pioneering studies have already shown the clinical applicability of these functional assays to guide treatment selection ^18–25^. For example, we have developed pharmacoscopy, a high-throughput *ex vivo* drug sensitivity screening platform with single-cell resolution, which showed promising potential to guide treatment selection leading to improved overall survival in AML patients ^18,19,23,26^. Few studies have started to address the intra-patient functional heterogeneity of diverse AML populations in response to *ex vivo* drug perturbations ^22,27^. However, it still remains uncertain if, and to what extent, drug testing across the cellular AML hierarchy improves clinical response predictions and functional precision medicine for AML.

## Results

### Multiplexed imaging and deep-learning of AML cell heterogeneity

To assess the functional impact of AML cell maturation heterogeneity on response to first line therapy (3+7, cytarabine plus daunorubicin), we collected a total of 174 samples from 44 newly diagnosed AML patients (Figure 1A, Table 1 and Table S1). Specifically, we prospectively collected bone marrow (BM) and peripheral blood samples at three different time points per patient: Prior to first-line induction chemotherapy, after hematopoietic recovery of the first and second induction cycle (Figure 1A & B). All samples were processed directly on the same day of biopsy and stained after 24 hours of incubation with drugs and control compounds. The patient cohort spanned a diverse set of WHO subclassifications (Table 1) and underlying molecular entities (Table S2). The median age of the study population was 57 years and the sex distribution was 64% male vs. 36% female. According to ELN 2022 risk categories ^28^, 20% of the patients were classified as favorable, 36% intermediate and 43% adverse. 77% of the patients achieved a complete remission after the second induction cycle of which 41% still had detectable minimal residual disease (Table 1 & Figure S1A).

**Figure 1.**
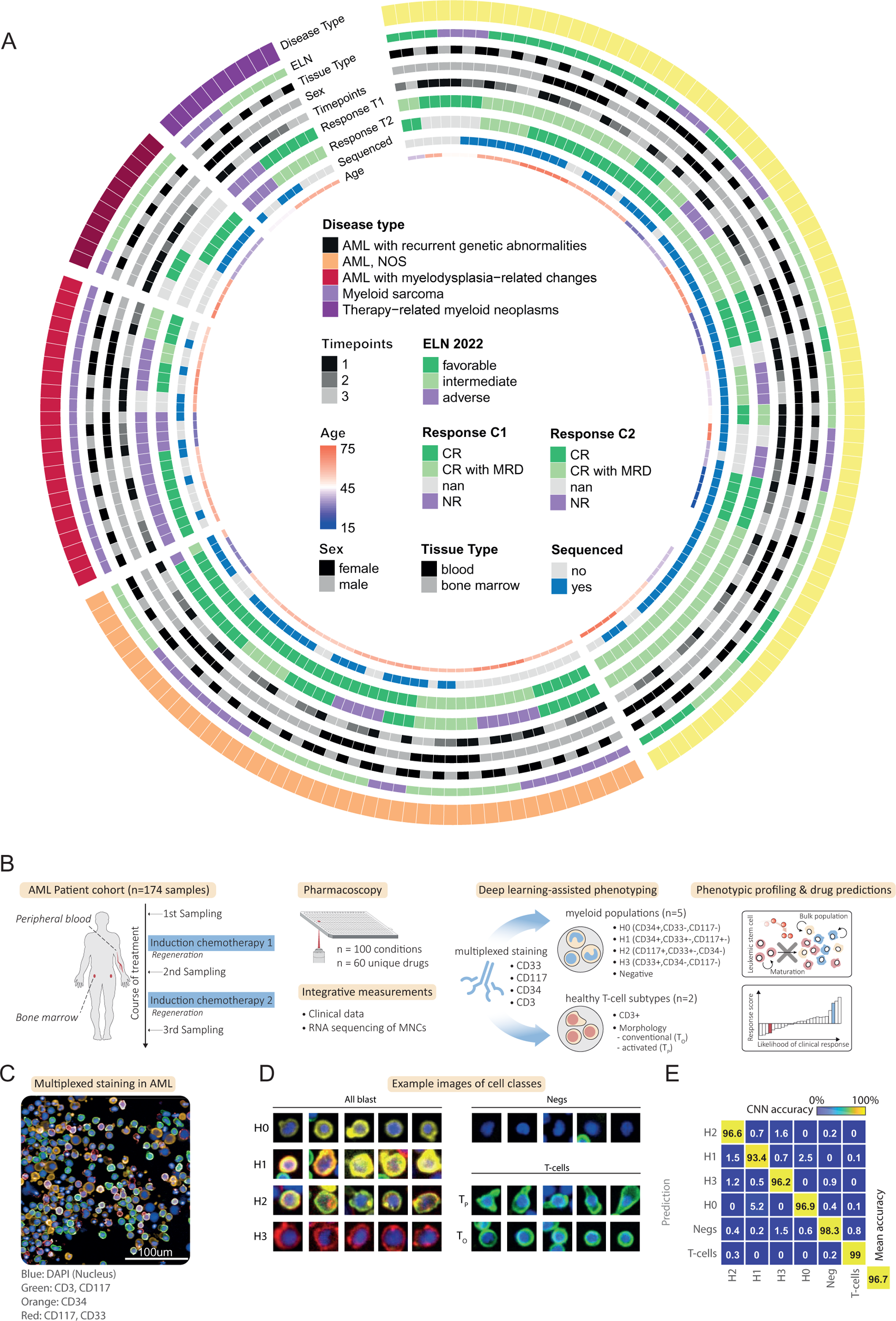
**A**, Circos plot overview of the newly diagnosed AML prospective patient cohort (174 samples from 44 patients). Concentric circles from outermost to innermost show the disease subtype, the ELN risk classification, the tissue type of each sample, patient sex, time point of sampling, clinical patient response after the first and second induction cycle, if a sample was sequenced and patient age at sampling. See Table 1 and Supplementary Table 1&2 for more cohort information. **B,** Workflow integrating *ex vivo* drug sensitivity screening (pharmacoscopy) with multiplexed single cell phenotyping, clinical data and direct molecular associations. Patient blood and bone marrow samples of newly diagnosed acute myeloid leukemia (AML) patients are taken over three time points and mononuclear cells are isolated and purified. Cells are seeded in 384-well plates, containing an indication-specific drug library. After 24h incubation, cells are fixed and stained with a multiplexed antibody panel and imaged by automated confocal microscopy. A deep learning pipeline consisting of several convolutional neural network (CNNs), classify each cell into debris, 4 cancer populations, T-cells or negative stained cells. T-cells get additionally evaluated for polarized or round morphologies indicating their activation status. Finally, *ex vivo* drug response prediction and a multi-omics analysis are calculated to identify optimal treatment options and clinically relevant subpopulations. **C,** Example image of multiplexed staining immunofluorescence on a AML sample at diagnosis. Green: CD3, CD117; Orange: CD34; Red: CD117, CD33; Blue: DAPI. Imaged at 20x resolution. **D,** Representative example images of curated cells for AML blast populations (CD34+ and CD33+ single positive cells (H0 and H3), combinations of CD33+/-CD117+ (H2) or CD33+CD34+ +/- CD117 (H1) marker positive cells, activated T-cells (T_P_) and conventional T-cells (T_O_) as well as cells negative for all stained markers (Negs). The size of crops is 48×48 pixels (14.4×14.4µm). **E,** Confusion matrix of CNN performance across 1000 test cells per class.

**Table 1.**
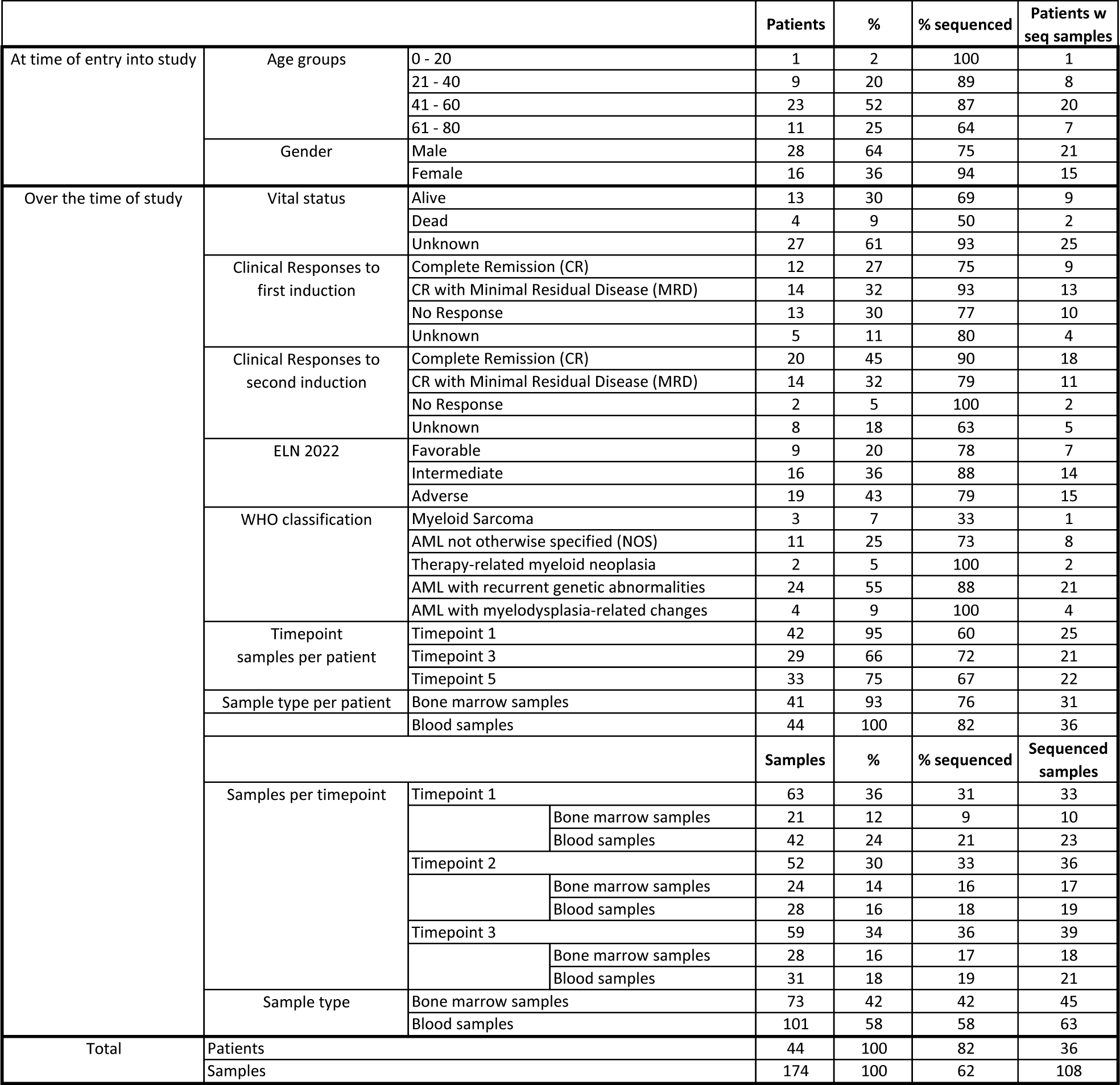

To characterize different functionally divergent AML subpopulations and associated non-malignant immune populations, we adapted our recently developed high-content multiplexed immunofluorescence approach ^29^. This new workflow enables the simultaneous integration of high-throughput drug screening with multiplexed single-cell phenotyping and associated molecular measurements in a single round of staining and imaging. To identify differentiation stages of AML blasts, we stained for the surface markers CD34, CD117 (c-kit) and CD33 (Figure 1C). CD34 is known to be expressed on undifferentiated AML blasts, including leukemic stem cells. CD117 is expressed on less differentiated AML blasts at different stages of early lineage commitment, while CD33 is present on more mature cells. As myeloid differentiation occurs, CD33 expression levels typically increase, while CD34 and CD117 expression decreases ^30,31^. In our multiplexed workflow, we additionally stained for CD3 to capture the T-cell population, furthermore allowing us to probe the inflammatory state via morphological T-cell profiling ^29,32^ and to assess off-target drug effects (Figure 1C).

We employed deep learning to identify putative AML cells and classify their maturation-associated marker profile. Specifically, we used a convolutional neural network with an adapted ResNet architecture (Figure S2A), as has been previously used in this context with high accuracy ^19,29^, to classify each cell into six mutually exclusive and marker-defined cell populations: Putative AML cell classes included CD34+ single-positive cells (referred to as AML class H0), CD34+ double-positive cells expressing CD33+ and/or CD117+ (AML class H1), CD34- and CD117+ cells optionally expressing CD33+ (AML class H2), and CD33+ single-positive cells (AML class H3). We furthermore classified CD3+ T-cells, and cells negative for all stained markers (Negs) (Figure 1D).

The Convolutional neural network (CNN) was trained and tested across 60,000 manually curated example images from across patients, samples, and timepoints, and achieved a classification accuracy of 96.7% in a ten-fold cross validation setting (Figure 1E & S2B, dataset available at: https://doi.org/10.3929/ethz-b-000680077). T-cells were additionally evaluated for distinct polarization and activation-associated phenotypes, as previously established (Figure S2C & 1D) ^29,32–34^. We further applied a ‘clean-up’ CNN analyzing DAPI and brightfield phenotypes, which was used to filter out dead cells, cell debris, segmentation errors, and other artifacts (Figure S2D). After clean-up, remaining cells showed limited levels of apoptosis in validation experiments (Figure S2E). We also included non-multiplexed control stains on each plate, allowing us to evaluate false-positive class assignments. This revealed no signs of systematic misclassification (Figure S2F) and highlighted the excellent technical reproducibility within control wells and across drug conditions (average correlation between replicates: 0.99) (Figure S3 A & B). Lastly, we analyzed the robustness of the approach by screening samples from three independent patients with both a 24- and 48-hour drug incubation. The longer incubation only showed minor impacts on baseline population abundances and associated drug screening results (Figure S3C).

To validate our image-based deep learning workflow, we compared the results with sample-matched flow cytometry performed as part of the clinical routine. Population comparisons were possible for 27 samples at diagnosis with matched CD34, CD117, and CD33 staining. We employed two complementary flow cytometry gating strategies: one replicating the CNN-matched marker-based populations on all cells, and the other analyzing marker expression specifically within the commonly used CD45dim blast populations (Figure S4A). At diagnosis, the vast majority of CD34, CD117, or CD33 expressing cells were part of the CD45dim blast gate (90%, 95%, and 92%, respectively), and only 3% of CD45dim blasts were not captured by these markers (Figure S4B). Furthermore, we observed high correlations between the cell population abundances quantified by our image-based approach and flow cytometry across the 27 patient samples (average Pearson correlation:0.61; Figure S4C & D). Thus, our image-based approach accurately captured the majority of AML cells present in these samples at diagnosis.

In summary, through combining single-cell microscopy of AML patient samples with multiplexed immunofluorescence and cell-based CNNs, we have developed an efficient method able to robustly and rapidly profile cellular phenotypes in high-throughput imaging of AML biopsies.

### Phenotypic profiling quantifies the AML cell hierarchy across patients

We next analyzed the heterogeneity in hierarchical profiles and clinical associations across the patient cohort. We observed a high level of heterogeneity in the AML cell-type composition of AML patient biopsies (Figure 2A): CD34 expressing populations (H0 + H1) showed an inverse correlation with CD33 (H3) (Figure S5A). CD34 expression (all CD34+) was associated with fewer myeloid marker-positive cells but more with marker-negative cells, whereas CD33 (H3) expression was strongly linked to the overall abundance of blasts (Figure S5A). To investigate the differential contributions of sampling time, tissue type, patient age and sex to population heterogeneity and marker expression variations, we conducted an analysis of variance (ANOVA) (Figure S5B). As expected, general subpopulation composition was mainly influenced by the sampling stage during first-line induction therapy, reflecting the response to treatment. Sampling bone marrow or peripheral blood showed no systematic influence on the cancer subpopulation composition, with only T-cell and marker-negative cells significantly differing between tissues. While patient sex did not influence population composition, age was associated significantly with tumor burden, with older patients having more AML cells relative to T-cells and marker negative cells, consistent with their known poorer prognosis (Figure S5B).

**Figure 2.**
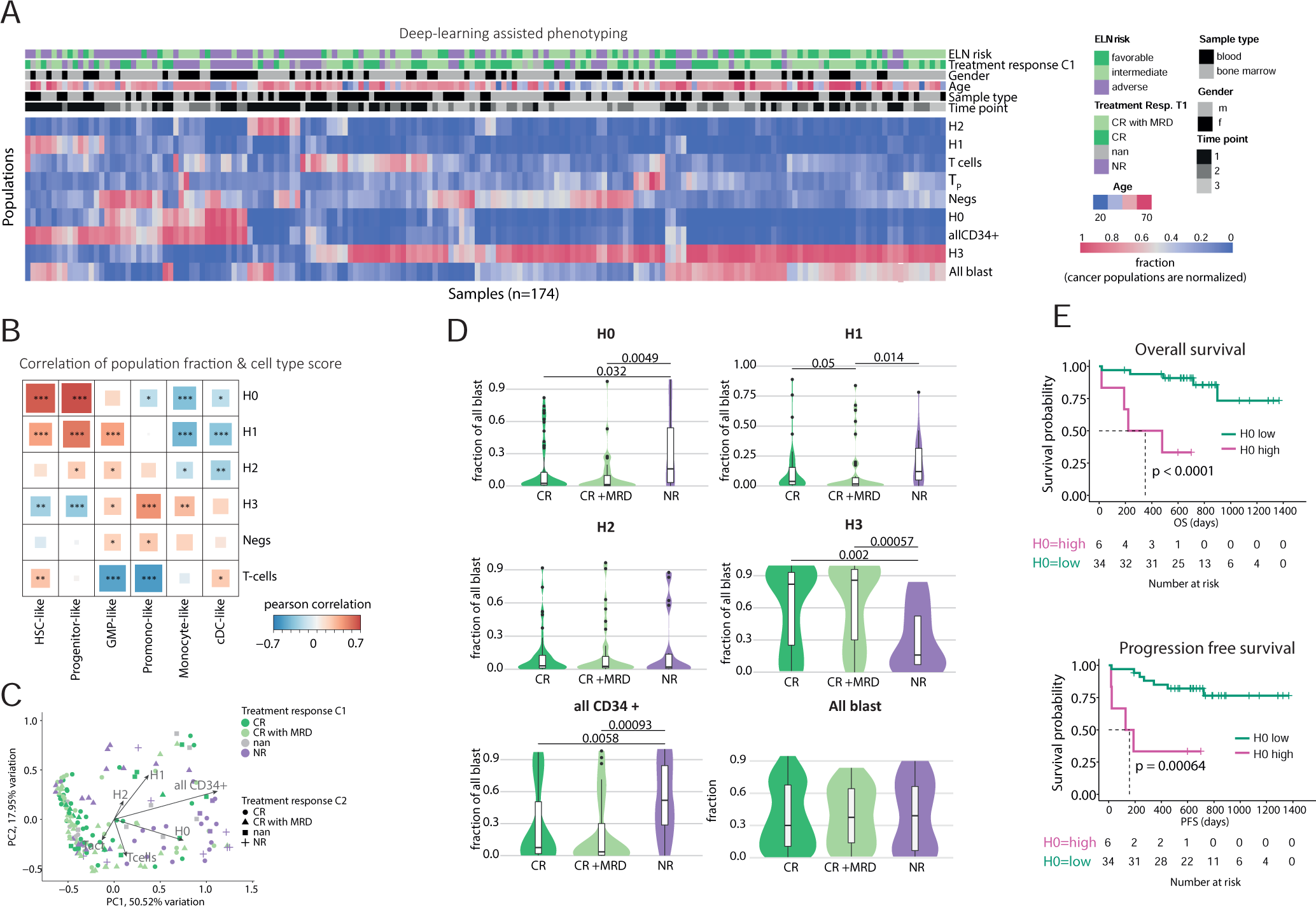
**A**, Deep learning-assisted phenotyping of all samples. Heatmap shows the mean population fractions across DMSO treated wells per sample. H0, H1, H2, H3, all CD34+ population fractions were normalized towards the absolute AML abundance. Samples are annotated with additional clinical annotations, as presented in the legend. **B,** Correlation of population abundance at diagnosis and AML cell type scores. AML cell types scores were calculated as defined by Van Galen et al. ^31^ and correlated to absolute population abundances for each sequenced sample. **C,** Principal component analysis of population fractions of all samples shown in A colored by clinical response to the first induction round. Shapes indicate response to the second induction round. **D,** Population abundance in relation to treatment outcome. Violin plots display the distributions of different population fractions measured before clinical treatment response (first or second round). CR=complete remission. MRD=minimal residual disease. NR=non responder. Samples with no response were excluded from analysis. Like in A, AML population fractions were normalized towards the absolute AML abundance. **E,** Blood CD34 single positive abundance (H0) at diagnosis-based stratification of progression-free (PFS) and overall survival (OS) (n=42 patients) plotted as Kaplan-Meier survival curves. Two patients were excluded because no sample at diagnosis was available. H0 high population is defined as an absolute fraction higher than 0.083. P Value asterisks are defined as following: *p<0.05, **p<0.01, ***p<10^-3^.

Van Galen et al. recently identified gene expression patterns for six distinct cell types in AML along their maturation axis using single-cell sequencing. This included hematopoietic stem cell (HSC)-like, progenitor-like, granulocyte-monocyte progenitor (GMP)-like, promonocyte-like, monocyte-like and conventional dendritic cell (cDC)-like types ^31^. To transcriptionally characterize our image-based AML maturation stages, we performed bulk RNA sequencing on 103 matched samples (Figure S6A-D), and correlated cell type scores, derived from the gene sets previously published ^31^, for each sample with the image-based sample composition (Figure 2B). Globally, the bulk transcriptome at diagnosis reflected the abundance of different maturation states: as anticipated, the fraction of CD34-expressing populations (H0 and H1) demonstrated the strongest correlation with the HSC and progenitor-like cell signatures. With the loss of CD34 expression, the population loses its transcriptional HSC association, and with decreased CD117 expression, the progenitor stage as well. The CD33+ H3 population was primarily associated with GMP, pro-mono and monocyte-like cell type associations (Figure 2B). Furthermore, we correlated the image-based subpopulation abundances with a previously reported transcriptional AML stemness signature, the LSC17 score ^11^. Again, CD34 single-positive cells (H0) exhibited a mild but significant positive correlation with the LSC17 stemness score, followed by the H1 population, while CD34-negative populations (H2 & H3) showed no significant association (Figure S6E). Thus, the image- and marker-based strategy classified AML maturation stages in line with previously reported transcriptional maturation signatures.

We next sought to investigate therapeutic resistance to drugs by investigating the known relationship between Tumor necrosis factor alpha (TNF-α) and the maintenance thus chemoresistance of LSCs ^35^. We therefore treated cells from three AML patients at diagnosis with increasing concentrations of TNF-α *in vitro*, and analyzed results by image-based profiling (Figure S7A). Following 48 hours of treatment, we observed a significant increase in the CD34+ (H0) abundance (Figure S7A) and a decrease in CD117CD33 (H2) cells. While prolonged ex-vivo exposure to cytarabine resulted in a significant reduction of the CD34+ population, the presence of high levels of TNF-α substantially increased their chemoresistance (Figure S7B), supporting the notion that CD34+ cells represent a stem cell-like population with the TNF-α pathway as a potential target in AML management.

Our combined findings therefore confirm that the identified populations represent distinct stages of AML blast differentiation. The most immature, stem cell-like H0 population is marked by CD34 single-positive cells, followed by the H1 population. The more differentiated H2 cells precede the most mature H3 CD33+ single-positive cells, illustrating a clear progression in AML blast maturation in our population definition (Figure 2B).

### AML cell maturation and sample composition predicts clinical outcome

LSCs hold significant prognostic importance due to their unique properties which render them resistant to standard treatments ^36^. We therefore investigated whether our image-based AML maturation profiling could stratify the clinical outcomes across the cohort. Utilizing principal component analysis of patient sample composition, we observed that patients were clustered according to their response to first-line treatment (Figure 2C). In line with previous reports, AML blast maturation was a strong clinical predictor of treatment response, progression-free survival and overall survival, whereby high abundance of CD34 expressing cells prior to treatment strongly predicted poor treatment response (Figure 2D-E and Figure S8A-C) ^6,8–10,36^. Furthermore, a high fraction of H0 stem cells in blood at diagnosis was a significant stratifier of progression free and overall survival, notably better than the ELN risk score within this cohort (Figure 2E and Figure S8D). For patients who achieved complete remission (CR) after the first round of treatment, we observed a significant relative reduction of cells with immature phenotypes. On the other hand, persistent immature cell phenotypes were observed in patients experiencing treatment failure to the first induction chemotherapy (Figure S9A). At the end of the second cycle, we noted an increase in H3 cells, in patients who achieved CR compared to patients which did not respond, likely indicating a recovery of the healthy hematopoietic environment (Figure S9B). Finally, we aimed to determine whether our findings could be replicated using standard diagnostic flow cytometry. While the correlation between the H0 population and our readout was modest (Figure S4C & D), the relative abundance of this population within all marker-positive groups prior to treatment initiation remained a significant predictor of first-line treatment response when assessed by flow cytometry (Figure S10).

Using a recently developed tool to profile T cell activation via morphological polarization ^29^, we investigated whether additional AML clinical parameters could be predicted. An interesting side-observation was that T-cell morphology appeared predictive of several AML relevant clinical parameters. Onset of neutropenic enterocolitis (NE), a critical complication that is associated with higher mortality, was associated with the inflammatory morphological signature of T-cells. Although patients which developed NE showed an overall higher amount of T-cells prior to treatment initiation, we observed that those T-cells showed a significantly lower inflammatory morphological signature compared to patients who did not develop NE (Figure S11A). Furthermore, in patients with remaining minimal residual disease (MRD), we observed a significant increase in the inflammatory T-cell subset compared to pre-treatment levels (Figure S11B). This persisted even after hematopoietic recovery following the second induction cycle, distinguishing MRD+ patients from those who achieved a CR (Figure S11C).

In summary, our high-content, image-based profiling of the AML cellular hierarchy effectively stratifies patients based on clinical outcomes including treatment response, neutropenic enterocolitis, progression-free survival, and overall survival. Importantly, CD34 single positive H0 abundance could serve as an accessible diagnostic marker for chemoresistance to cytarabine and anthracycline-based first-line induction treatment.

### AML drug response profiles are mainly determined by the degree of maturation

To gain deeper insights into intra-patient drug response heterogeneity in relation to AML maturation stages, we screened a set of 80 unique drugs and selected drug combinations (Figure 3A & Supplementary Table 3). This drug panel includes not only common first-line AML treatments but also a variety of FDA-approved blood cancer drugs, such as tyrosine kinase and immune checkpoint inhibitors. To quantify chemo sensitivities across the cellular maturation hierarchy, we calculated relative blast fractions (RBFs) as previously described and clinically validated ^18,19,23,26^, for all four AML cell maturation classes within a sample. Here, positive scores indicate a higher chemosensitivity of this population compared to others, whilst negative scores denote relative *ex vivo* chemoresistance. Clustering the complete profiles revealed substantial drug response heterogeneity among cancer subpopulations and patient samples (Figure 3A), with the drug response of an AML blast population being strongly influenced by its maturation stage (Figure B). Notably, within-sample drug response differences following the degree of maturation strongly outweighed between-sample drug response differences (Figure 3). Furthermore, different drugs were strongly clustered by mode-of-action and class, demonstrating a high degree of technical robustness of this dataset and enabling mode-of-action investigation of less well characterized drugs (Figure 3A & B).

**Figure 3.**
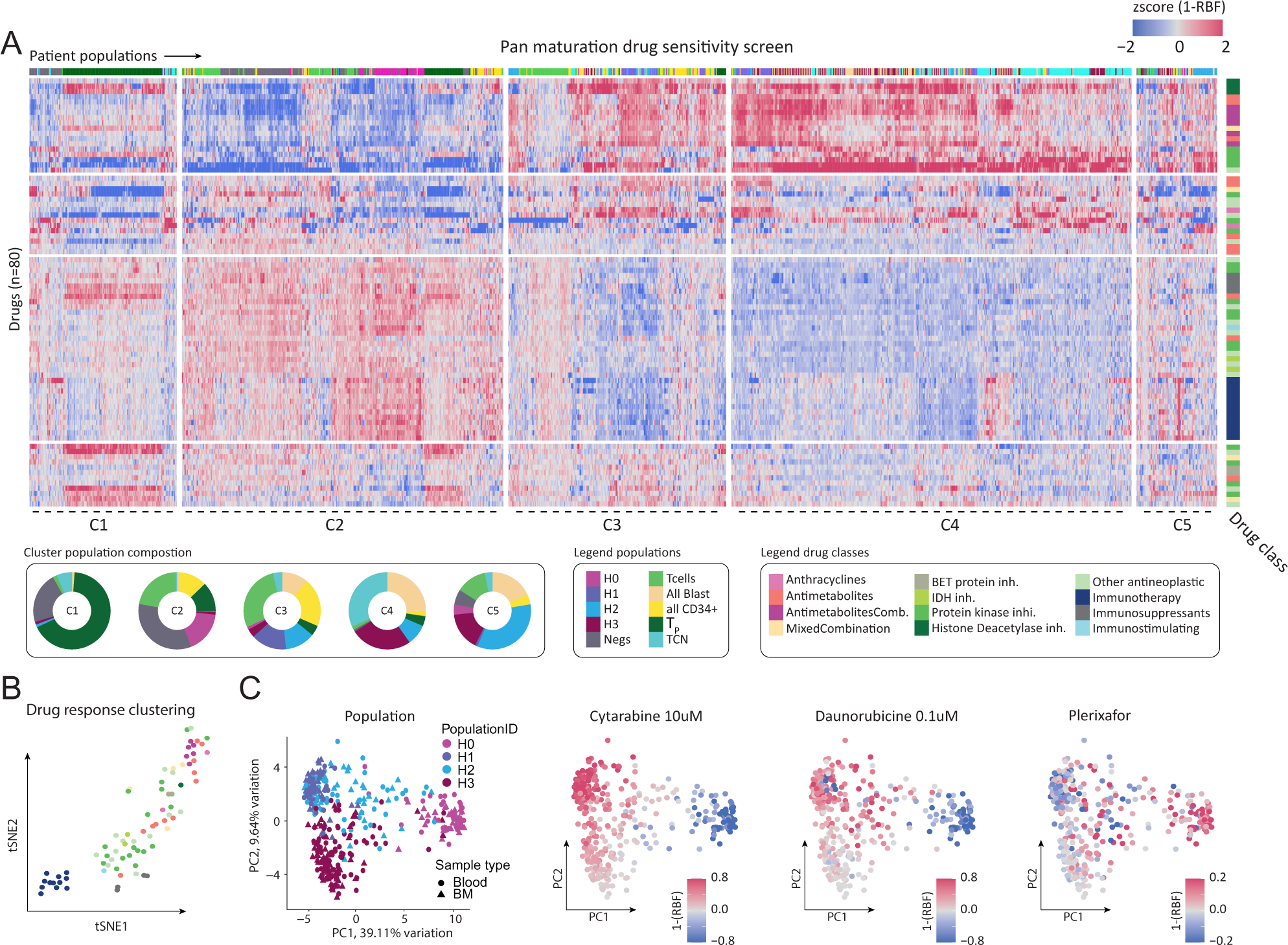
**A**, Overview of the pan-maturation drug screen. Individual data points represent relative blast fraction scores (1-RBF) of a sample specific population per single drug. Low abundant populations (fraction below 0.05) were excluded. Values were standardized (z-scored) within each treatment (n=80). Columns of the heatmap were grouped into five individual clusters (C1-C5) and the donut plots show the proportion of populations within each group. **B,** t-Distributed Stochastic Neighbor Embedding (t-SNE) visualization of all drug responses shown in A per individual tested treatment. **C,** Principal component analysis of AML population drug responses of all samples colored either by population ID or drug response to cytarabine, daunorubicin or plerixafor.

To estimate a drug’s general effectiveness against different blast populations, we calculated the fraction of patient samples displaying a significant on-target effect across all samples for each population (Figure S12A&B). While the more mature H1, H2 and H3 populations exhibited high sensitivities to common first-line treatments across the majority of samples, such as anthracyclines, anti-metabolites, or selected protein kinase inhibitors, stem cell-like H0 cells generally displayed ex-vivo chemoresistance, including to first-line treatments cytarabine and daunorubicin (Figure 3C & S12A&B). We thus probed the dataset for drugs specifically targeting the CD34 LSC-like cells. The only compounds with limited general activity against the CD34 population were Plerixafor (relative chemosensitive in 21% of H0 populations) and dexamethasone (relative chemosensitive in 24% of H0 populations) ^37^. Dexamethasone (Dex) is a synthetic corticosteroid medication with potent immunosuppressive properties, and is often used in supportive care during AML treatment. Intriguingly, both treatments have been shown to exhibit activity against CXCR4 ^37–39^, a chemokine critical for the maintenance of leukemia cells within the bone marrow. Although we did not identify a universally effective treatment against the H0 stem cell compartment, we found at least one drug per patient that the H0 population was sensitive to ex vivo (defined by an absolute reduction of the population) for the majority (65%) of patients.

We further investigated the differential impact of patient age, sampling time point, and sampling location (tissue) on drug response for each population by ANOVA (Figure S13A). Elderly patients, who are known to have a lower tolerability to first-line induction chemotherapy, showed a significantly reduced *ex vivo* response to nucleoside analogs cytarabine, cladribine, clofarabine, and fludarabine (Figure S13B&C). In contrast, the immunomodulator pomalidomide demonstrated improved on-target scores in elderly patients, potentially providing a better-tolerated treatment regimen for this demographic (Figure S13B). However, both pomalidomide and lenalidomide exhibited significant compartment-dependent differences, with weakened responses against cells from bone marrow compared to blood (Figure S13D &E). In contrast, gilteritinib showed stronger on-target responses in bone marrow samples, possibly indicating a higher sensitivity to FLT3 inhibition in this tissue context (Figure S13D).

Lastly, we evaluated to what degree drugs activated T-cells *ex vivo*. Venetoclax was recently shown to directly activate T-cells and enhance T-cell-mediated antileukemic activity ^40^. Consistently, in our screen, venetoclax also emerged as one of the top activators of T-cell activity but surprisingly midostaurin and kinase inhibitors alvocidib and crenolanib emerged as very potent activators of T-cell activity, revealing a potential new anticancer mechanism of these drugs (Figure S13F).

In summary, we systematically analyzed the drug response heterogeneity and sensitivity profiles of various AML maturation stages, revealing maturation to be an extremely strong determinant of drug sensitivity. This high dimensional dataset, featuring differential population, time and tissue resolution will be made accessible to the community.

### Predictive power of pharmacoscopy for clinical response in newly diagnosed AML and identification of personalized treatments

We next evaluated the power of pharmacoscopy-based drug testing to predict clinical response of newly diagnosed AML patients to their first line treatment. To account for the vast heterogeneity observed across donors, we developed a ‘mean maturation score’ (MMS), which is calculated as the average response of each AML subpopulation as well as the all marker positive class and the total cell number readout (Figure 4A & S14A). Drug responses were included into the MMS only for AML subpopulations that were present in at least 0.5% of all nucleated cells in a sample in control conditions. The MMS score also considers the unwanted reduction of the T-cell population, thereby penalizing off-target cytotoxicity.

**Figure 4.**
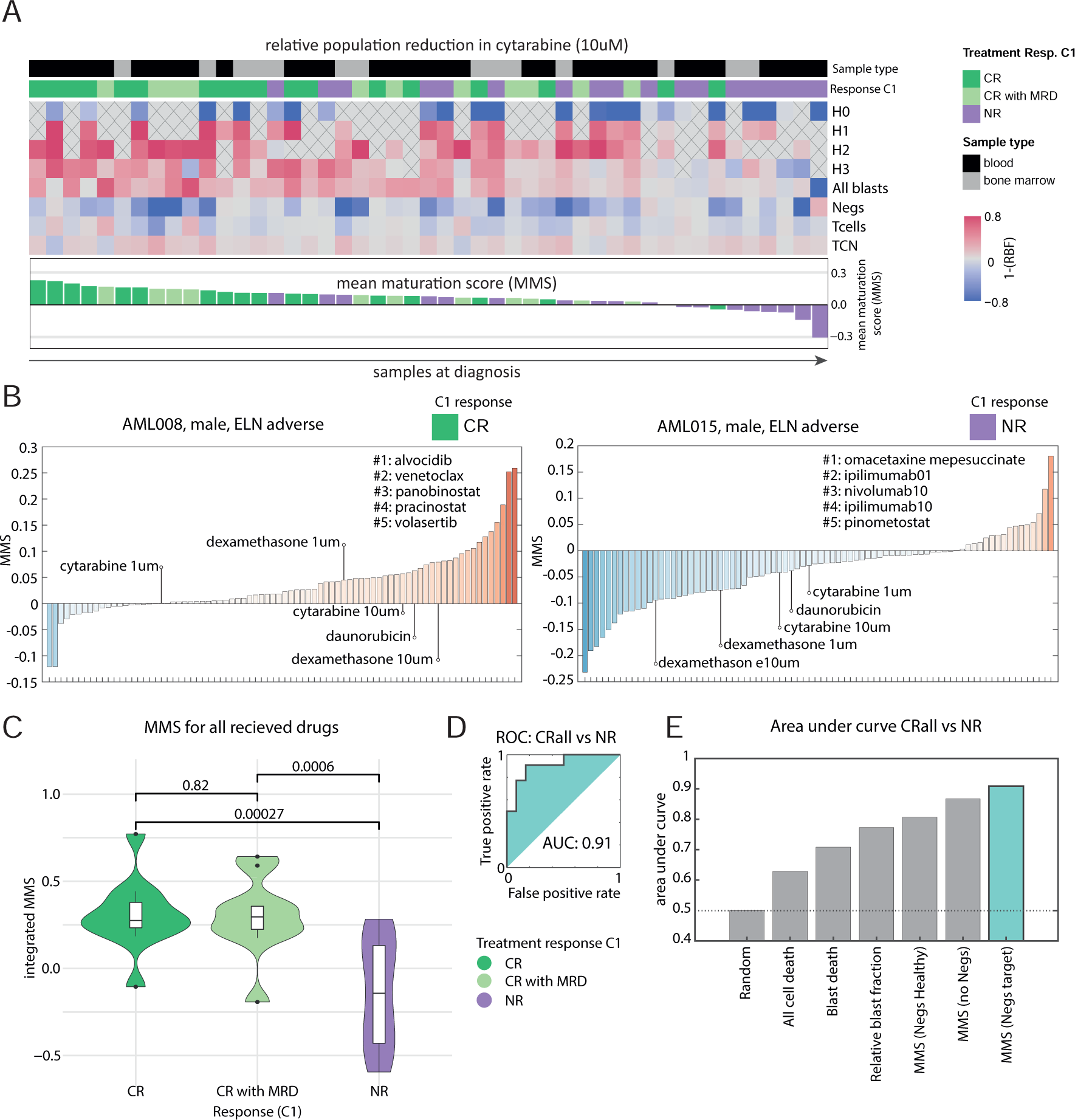
**A**, Cytarabine response at diagnosis. Top: Heatmap shows cytarabine response (1-RBF) for all samples at diagnosis split by population. Responses for low abundant populations (fraction below 0.05) are plotted in gray and crossed out. Columns of heatmap are sorted by the average response calculated across all populations, referred to as the mean maturation score (MMS), for more details see methods. Bottom: Sample MMS scores plotted as matching bar graphs. **B,** MMS all screened drugs for two selected patients, with corresponding patient numbers, clinical outcome, timepoint and screened tissue. Bar graphs show drugs ranked by the MMS. **C,** Violin plot shows the integrated MMS at diagnosis stratified by treatment response (n=34). Integrated MMS score was calculated by summing all MMS scores for every individual chemotherapy a patient was treated with. If multiple tissues (blood and bone marrow) were measured per patient, the average score across both samples was calculated per patient. Measurements were taken before a patient was treated. Patients without T1 samples (n=2), clinical response annotation (n=5) or patients which received drugs not in the screening panel (n=3) were excluded from this analysis. CR=complete remission. MRD=minimal residual disease. NR=non responder. **D,** Receiver operating characteristic (ROC) curve plot based on the integrated AUC values from responding and non-responding patients shown in A. The ROC curve shows the trade-off between sensitivity and specificity for the optimal MMS cutoff stratifying responders from non-responders. The area under the curve (AUC) is 0.91. **E,** Comparison of AUC rankings for different drug response readouts. AUC values were calculated based on ROC curves generated from different integrated drug response measurements of responding and non-responding patients. The AUC values ranged from 0.5 to 0.92, with the highest AUC value indicating the most accurate drug response readout in distinguishing between responding and non-responding patients.

As an example, Patient AML008 demonstrated significant on-target *ex vivo* effects for a broad panel of antineoplastic agents, including first-line treatments cytarabine and daunorubicin (Figure 4B left). Subsequent treatment with the standard ‘7+3’ regimen led to a complete remission for this patient. In contrast, Patient AML015 showed strong off-target effects for both agents *ex vivo*, and subsequently no clinical response to the chemotherapy combination of cytarabine and daunorubicin (Figure 4B, right). However, *ex vivo* drug testing identified sensitivity to both the protein translation inhibitor omacetaxine and immune checkpoint inhibition, suggesting an alternative personalized treatment regimen might have yielded better clinical outcomes for this patient. To evaluate the predictive power of pharmacoscopy in our whole AML cohort, we integrated the MMS drug responses for each individual chemotherapy treatment a patient received (on average 3.8 drugs per patient) (Figure 4C). We next related the response to the first induction round with the average integrated MMS response across both patient bone marrow and blood samples at diagnosis. The integrated drug sensitivity score successfully distinguished non-responders from responders with an area under the ROC curve of 0.91 (Figure 4C&D, S13A), highlighting the predictive power of our *ex vivo* functional drug testing platform. Furthermore, systematic comparison of the AUCs for different drug response readouts showed that considering the cellular AML hierarchy is the most accurate pharmacoscopy-based response prediction for this cohort, while bulk cell death performed barely above random (Figure 4E). Interestingly, we observed that the predictive accuracy of our readout improved when we also considered the drug responses of the marker-negative population as a target, indicating the presence of potential malignant cells or other cancer-associated cells within that class (Figure 4E).

In light of the accurate predictions of first-line treatment response, we were interested in determining if we could identify alternative treatments that might have been effective for non-responding patients (n=13; NR to 1st cycle). While cytarabine had a minimal MMS for these patients, we could additionally identify on-target treatments for every non-responding patient. Here, the most effective individual agents identified in our screen demonstrated significantly higher on-target MMS values than cytarabine (Figure S14B). These individual compounds varied considerably between patients, and included omacetaxine, panobinostat, ipilimumab, a combination of cytarabine and fludarabine, and alvocidib (Figure S14C). Furthermore, we ranked the drugs based on their effectiveness per patient at diagnosis to identify treatments that consistently ranked highly across the entire cohort (Figure S14D). In this analysis, alvocidib emerged as a top-performing drug with favorable *ex vivo* drug scores. Alvocidib is a CDK9 kinase inhibitor ^41^ and currently in clinical trials with initial promising results and could provide an additional complementary candidate for more effective first-line treatments ^42^. Also Omacetaxine, cladribine, clofarabine, crenolanib, venetoclax, and azacytidine demonstrated promising *ex vivo* drug rankings across the donor cohort (Figure S14D). Interestingly, Venetoclax is already in use in combination with other agents, such as the hypomethylating agent azacitidine, which led to improved response rates compared to previous regimens ^43,44^. In conclusion, our study showcases the potential of pharmacoscopy in predicting treatment response in AML patients, allowing for more personalized approaches to therapy. By taking into account the targeted reduction of the entire AML cellular hierarchy, our method accurately distinguishes between responders and non-responders in a prospective observational setting. The identification of alternative treatments that might have been effective for non-responding patients highlights the potential for optimizing treatment strategies for AML patients, ultimately improving clinical outcomes. With agents such as alvocidib and venetoclax demonstrating promising results, there is potential for developing more effective first-line treatment options for AML patients in the future.

### Improving response prediction and treatment selection in resistant/relapsed AML

Lastly, we aimed to validate our findings in a relapsed/refractory (r/r) AML setting. We recently established that pharmacoscopy can be employed for therapy selection in rr-AML patients with no registered treatment options and demonstrated clinical benefit in heavily pretreated and frail patients ^23^. We now evaluated whether we could retrospectively improve response predictions for this rr-AML cohort in a post hoc analysis. Most patient samples were analyzed by pharmacoscopy utilizing a CD33, CD34 & CD3 antibody staining panel and we therefore attempted to define relevant AML hierarchy populations based on combinations of the different blast markers used. Similar to the drug response calculation in the newly diagnosed AML cohort, we then calculated the average response scores across all present populations.

In this analysis a relative reduction of marker-positive cancer cells across all subsequently received treatments showed promising but not statistically significant trends. On the other hand, the integrated mean maturation score (MMS) significantly stratified patients achieving complete remission from those with progressive disease (Figure S15A). In contrary to newly diagnosed AML, we realized that in this r/r AML cohort, very small populations were less meaningful, and the most optimal separation was achieved by taking into account only more abundant AML subpopulations as well as the all marker-positive cell drug responses and the total cell number drug responses (Figure S15B). These findings highlight the importance of considering functionally different populations also in an r/r AML setting, as it has a significant impact on response predictions. In light of our findings, we highlight the case of patient P31 who received a pharmacoscopy-guided treatment with Cladribine, Cytarabine, and Venetoclax, and achieved a complete remission. Although cytarabine and cladribine induced a reduction in the overall cancer marker-positive population, including the CD33-expressing population, they did not effectively target the CD34 population. In contrast, venetoclax demonstrated strong ex vivo results against this specific population, emphasizing the importance of subpopulation resolution in personalized treatment selection (Figure S15C).

## Discussion

Although the diagnostic significance of AML blast maturation is becoming increasingly apparent, it is not yet considered in personalized treatment recommendations. In this study, we developed a high-dimensional single-cell drug testing platform as a new tool to screen and investigate differential chemoresistance of functionally diverse AML populations within patient samples. In line with recent research ^22,27^, we demonstrated that the AML cell differentiation spectrum, specifically leukemic stem cell-like H0, influences patient progression and drug sensitivity. While LSCs are typically defined as CD34+ and CD38-cells, a marker combination we did not assess, our findings demonstrated that our H0 CD34+ CD117-CD33-population definition exhibits gene expression signatures and functional behavior similar as those described for leukemic stem cells. Our results are thus in line with previous literature regarding the negative prognostic significance of leukemic stem cells and the CD34 marker. Consequently, we propose the abundance of the CD34 single-positive population as an accessible diagnostic marker for chemoresistance to cytarabine and anthracycline treatment, which can also be detected using standard diagnostic flow cytometry.

These results highlighted the efficacy of our screening technology in simultaneously evaluating drug effects on functionally distinct and clinically meaningful populations. This allows us to achieve several objectives: First, our platform offers a unique tool for high-dimensional characterization of drug effects in AML. Common drug testing tools either assess drug effects on AML cell lines (distant from patients) or examine drug impacts in a bulk fashion on all cells. In contrast, our technology offers a deeper insight into the mode of action of tested compounds, including their potential to target small but relevant populations or activate healthy immune cells. For instance, our screen revealed partial activity of dexamethasone against LSCs, an effect which would be hidden by bulk measurements. Additionally, we demonstrated that common AML treatments like midostaurin have previously unknown effects on the T-cell compartment, revealing new anti-cancer mechanisms.

Second, by integrating the response of AML in different maturation stages our technology improves response predictions and allows for better treatment recommendations. These findings underscore the importance of considering maturation stages in newly diagnosed AML for personalized therapeutic strategies and optimal patient outcomes, highlighting the need for increased attention in future studies and personalized treatment plan development for AML patients. We also observed the positive impact of assessing functionally distinct populations in an independent relapsed AML cohort. Here, however, addressing very small populations diminished prediction accuracies, in contrast to newly diagnosed AML, where even small populations favored positive prediction accuracies. This discrepancy could be explained by a strong clonal evolution induced by multiple treatment rounds experienced by relapsed AML patients which could disrupt maturation hierarchies. Nevertheless, drug response prediction improved when considering more than one functionally diverse population, supporting the concept. To evaluate this, future studies could monitor patients over time to determine whether the maturation spectrum changes and how this relates to functional responses across patients.

Lastly, our technology enables the recommendation of personalized treatment plans for each patient. Alvocidib emerged as the most generally effective treatment. Alvocidib is currently in clinical trials with initial promising results, and future results in larger cohorts will determine if it serves as a more general first-line treatment option than cytarabine or anthracyclines.

Despite the prospective character of this non-interventional study, limitations include the relatively small patient cohort, which may not capture the full spectrum of AML heterogeneity, encompassing various maturation patterns. The small cohort resulted in a limited number of non-responders after the second induction cycle, restricting robust statistical analysis and focusing our attention primarily on samples at diagnosis and after response to cycle 1. Future studies with larger cohorts would enable associations between specific mutations, chromosomal aberrations, and single-cell drug response patterns, providing valuable insights.

In conclusion, our high-dimensional single-cell drug testing platform offers a promising approach to address the challenges of AML heterogeneity and optimize personalized treatment strategies. Our findings underscore the need for greater attention to maturation stages in the development of personalized treatment plans for AML patients, with the goal of improving patient care and outcomes. Further research involving larger patient cohorts and longitudinal tracking of maturation spectrum changes would provide additional insights and refine our understanding of AML blast maturation’s role in personalized medicine.

## Methods

### Study design and participants

For this prospective, non-interventional, single-center study, samples and clinical data were collected from patients with newly diagnosed acute myeloid leukemias undergoing intensive induction chemotherapy. The samples were taken at three timepoints: prior to induction, after regeneration of the first chemotherapy cycle and after hematopoietic recovery following the two cycles. Patients were eligible for inclusion if newly diagnosed or with relapsed AML who have not been given systemic chemotherapy within the past 12 months prior to inclusion; the patient was over 18 years of age; the patient was receiving intensive induction chemotherapy; the patient was able and willing to provide written informed consent and to comply with the study protocol procedures. The recruitment of patients occurred through the project leader or the team of treating physicians of the Department of Medical Oncology and Hematology of the University Hospital Zurich (USZ) during daily clinical practice, when patients were hospitalized for diagnostic leukemia work-up and initiation of induction chemotherapy. Inclusion of patients strictly occurred before the start of induction chemotherapy. Clinical responses were measured by the remission state in the bone marrow sampled after regeneration of the hematopoietic system (about 28 days after start of chemotherapy). Here, blast persistence resulting from bone marrow cytology and histology were used as indication of treatment response. If patients displayed persistent blasts in the bone marrow at the prior aplasia control (a bone marrow puncture about 14-16 days after start of chemotherapy), a second induction cycle was induced without waiting for hematological recovery, resulting in no response data. Furthermore, the lack of response data in other incidences is a result of patients dropping out of or discontinuing in this study. The research project was carried out in accordance with the research plan and with principles enunciated in the current version of the Declaration of Helsinki (DoH), the Principles of Good Clinical Practice (GCP), the Swiss Law and Swiss regulatory authority’s requirements as applicable. Ethical approval was granted by the Ethics Committee of the Kanton Zurich (CEC Zurich, BASEC-Nr: 2018-01547).

### Pharmacoscopy

Bone marrow aspirate or peripheral blood tubes were provided by the team of treating physicians of the Department of Medical Oncology and Hematology of the University Hospital Zurich. To isolate MNCs, the samples were diluted in PBS (Gibco) + 2 mM EDTA (Sigma-Aldrich) and were purified using a Lymphoprep density gradient (STEMCELL Technologies) according to the manufacturer’s instructions. The resulting MNCs at the interface were collected, washed once with PBS+EDTA and resuspended in RPMI 1640 + GlutaMax medium (Gibco) supplemented with 10 % human serum (Chemie Brunschwig). The single-cell suspension of immune cells were seeded (2*10^4^ cell/well with 50 μl/well) in CellCarrier 384 Ultra, clear-bottom, tissue-culture-treated plates (PerkinElmer) containing antineoplastic agents (see Supplementary Table 3) and incubated overnight (24 h at 37 °C and 5 % CO_2_). Cell number and viability was determined by use of a Countess II Cell Counter (Thermo Fisher).

For all the patients, the drug screen library included 80 single drugs and drug combinations with two to six technical replicates in total (see Supplementary Table 3). The assay was stopped by fixing and permeabilizing the cells with 20 μl/well of a solution containing 0.5 % (w/v) Formaldehyde (Sigma-Aldrich), 0.05 % (v/v) Triton X-100 (Sigma-Aldrich), 10 mM Sodium(meta)periodate (Sigma-Aldrich) and 75 mM L-Lysine monohydrochloride (Sigma-Aldrich). After 20 minutes incubation at room temperature (RT), the fixative-containing media was aspirated by use of a HydroSpeed plate washer (Tecan). The cells were then blocked (50 μl/well) with PBS supplemented with 5 % fetal bovine serum (FBS, Gibco) and photobleached for 4 h to 24 h (at 4 °C) to reduce background fluorescence by illuminating the fixed cells with conventional white light LED panels.

### Multiplexed immunostaining and imaging

All fluorescent primary antibodies used to identify the target blast population were used at a dilution of 1:300 in PBS and included CD3, CD117, CD33, CD34 (see Supplementary Table 4). For nuclear detection, all antibody cocktails contained 2 µg/mL DAPI (4’,6-Diamidino-2-Phenylindole, Biolegend). Before the immunofluorescence staining, the blocking solution was removed, 20 µl/well of the antibody cocktail added and the imaging plate was incubated for 1 h in the dark (at RT). Thereafter, the staining solution was removed and PBS was added (70 µL/well). Each plate contained both multiplexed and staining control wells, the latter of which, located at each of the corners of the plate, had a reduced number of antibodies to serve the evaluation of antibody functionality and generation of the CNN-training data (see below). The 384-well plates were imaged by use of a PerkinElmer Opera Phenix High-Content automated spinning-disk confocal microscope. At 20x magnification with 5×5 non-overlapping images, covering the whole well surface, each well of the plate was imaged. The images were taken respectively from the brightfield (650-760 nm), DAPI/Nuclear signal (435-480 nm), GFP/FITC/Green signal (500-550 nm), PE/Orange signal (570-630 nm) and APC/Red signal (650-760 nm) channels. For further analysis, the raw .tiff images from the microscope were used.

### Casp3 experiment

To examine cell viability on the grounds of apoptosis, additionally isolated peripheral blood mononuclear cells (PBMCs) from three samples were used (see Figure S2E). For a full description on the workflow for this experiment see paragraph “Pharmacoscopy” above. Immunostaining and imaging were performed as described above (paragraph “Multiplexed immunostaining and imaging”). Here, the staining panel for two of the samples (AML046 blood T1 and AML051 blood T1) included CD117, CD33, Cleaved Caspase 3 and CD34, while the other (AML044 blood T1) was stained with CD38, CD34 and Cleaved Caspase 3 (see Supplementary Table 3). Cells were identified by maximum correlation thresholding on the DAPI channel as described below in more detail.

### TNF-α experiment

A treatment with TNF-α (Peprotech) and cytarabine (Sigma-Aldrich) was used for the induction and suppression of the CD34 single positive phenotype (H0). This experiment was performed on previously isolated (for a full description on the workflow see paragraph “Pharmacoscopy” above) PBMCs of three AML patients at diagnosis that were frozen as cell suspension with 10 % dimethyl sulfoxide (DMSO) upon isolation and stored at -80 °C. After thawing the samples, approximately 10*10^6^ cells were recovered, washed with RPMI 1640 + GlutaMax medium (Gibco) supplemented with 10 % human serum (Chemie Brunschwig), seeded (2*10^4^ cells/well with 50 μl/well) in duplicate in CellCarrier 384 Ultra, clear-bottom, tissue-culture-treated plates (PerkinElmer) and incubated for 48 h at 37 °C and 5 % CO_2_. Cell number and viability was determined by use of a Countess II Cell Counter (Thermo Fisher). The 384-well plates contained an increasing concentration of TNF-α (1 ng, 10 ng, 100 ng), cytarabine in 10 µM concentration and a combination (TNF-α in 100 ng; cytarabine in 10 µM) of such, each in two technical replicates (resulting in four technical replicates per sample per condition). As control for TNF-α treatment, PBS was used, while DMSO acted as control for the cytarabine treatment. Following incubation, the plates were fixed, blocked, stained and imaged as described in detail above (see paragraphs “Pharmacoscopy” and “Multiplexed immunostaining and imaging”).

### RNA sequencing

For RNA sequencing, approximately 2*10^7^ cells were washed with PBS+EDTA prior to resuspension and a 10 minute incubation (at RT) in 1 x RBC lysis buffer (Biolegend). The resulting RBC-free immune cells were then washed once in PBS+EDTA prior to being split in two technical replicates. The cells of the two replicates were each resuspended in 350 µl TRIzol reagent (Invitrogen) and then frozen (at -80 °C) until further processing. RNA extraction was performed with a Quick-RNA MiniPrep Kit by Zymo according to the manufacturer’s instructions. RNA sequencing was performed by the Functional Genomics Center Zurich (FGCZ). In brief, cDNA libraries were obtained as described here: ^45^. Illumina library was obtained via tagmentation using Illumina Nextera Kit. All samples were sequenced in a single run on a NovaSeq6000 (single read, 100bp, depth 20 Mio reads per sample). Illumina adapters, sequences of poor quality as well as polyA and polyT sequences were removed from the raw reads using TrimGalore v.0.6.0 with cutadapt v.2.0 prior to alignment. Reads were then aligned to the human reference genome GRCh38, v93 (Ensembl) using STAR v. 2.5.3a. Read per genes were counted with featureCounts v1.5.0. Gene counts below a threshold of 20 raw counts were filtered and raw counts were normalized (DESeq2 ^46^). Only transcripts annotated as ‘protein coding’ or ‘long non-coding RNA’ were considered in the subsequent analysis. In total 108 samples were sequenced (of which three samples are batch controls). Samples with more than 5% of counts assigned to hemoglobin and no detectable blasts were excluded (n=6).

### Cell detection and single cell feature extraction

CellProfiler v2 ^47^ was used for single-cell image analysis. Single cell detection and nuclear segmentation was performed by maximum correlation thresholding on the DAPI channel. To extract cytoplasmic measurement, cellular outlines were estimated by a circular expansion of 8 pixels around the nucleus. To measure the local intensity background around each single cell, a second larger expansion of 30 pixels was performed. Standard CellProfiler raw fluorescent intensities were extracted, log_10_ transformed and normalized towards the local cellular background as described in Vladimer et. al., ^48^.

### CNN dataset generation and normalization

The curated dataset used for the 6 class cancer CNN was generated as described in ^29^. In brief, 48×48 pixel wide single cell subimages were generated across all five measured channels. Single cell images were then manually curated for their respective class using custom Matlab scripts and included cells from all patients, both tissues and three timepoints. In total a dataset of 62318 hand selected images were generated. All intensity values as input for the 6-class Resnet were log_10_ transformed.

The T-cell morphology CNN dataset was generated in the same manner, creating a total of 3545 (20078 all) T_ACT_ cells and 10596 (65918 all) T_CON_ cells.

### Convolutional neural network (CNN) architecture, training and data augmentation

A 6-class 71-layer deep convolutional neural network with an adapted ResNet architecture (Figure S2A) ^49^ was implemented using MATLAB’s Neural Network Toolbox of Version R2021a. 48×48 pixel and 5 channel input images were used. The complete training and test dataset contained a total of 60000 cells with 10000 cells per class. To evaluate CNN performance the full dataset was randomly split into a training set and a test set (Test data with 10% of data; 1000 cells per class). The 6-class CNN was trained using randomly initialized weights and biases and the adaptive learning rate optimization ‘ADAM’. The network was trained for 20 epochs with an initial learning rate of 0.001 which was dropped every 5 epochs with a factor of 0.1. Furthermore, a mini batch size of 512 images and L2 regularization with 0.001 was applied. To further strengthen generalization, input images were augmented in each iteration. Here, images were randomly rotated in 45-degree steps with an additional possibility to be also flipped vertically or horizontally.

### T-cell polarization

The T-cell morphology CNN was trained to classify fixed and permeabilized human T-cells as T_P_ or T_O_ cells via DAPI, Brightfield and CD3 48×48 pixel (14.4µm x 14.4µm) single cell crops. The training set comprised 3301 (16777 all) T_P_ and 9805 (56113 all) T_O_ cells, derived from primary human samples. Testing was performed on 244 (1363 all) T_P_ and 791 (4638 all) T_O_ cells (approximately 10% of the training set). The network employed a 39-layer CNN based on an adapted ResNet architecture. Training was performed using the same parameters as described for the 6-class network. The network was trained five times to assess training robustness, and the most accurate network was selected for downstream analysis. These CNNs were used to classify individual T-cell crops into their output classes.

The fraction of T_P_ cells was normalized towards the absolute fraction of T-cells to represent the relative polarization of T-cells.

### Quality control and cellular cleanup

For quality control, we applied a custom small ‘Cleanup CNN’, which evaluated each individual cell for correct segmentation, contaminants or artifacts. Also here an adapted ResNet architecture was used (Figure S2A). The cleanup CNN was trained for 30 epochs, with a minibatch size of 512 images utilizing adaptive learning rate optimization ‘ADAM’. The training was started with an initial learning rate of 0.001 which was dropped every 5 epochs with a factor of 0.1. L2 regularization of 0.001 was employed. The hand-curated training and validation dataset (10% of the data) consisted of 60000 individual, 48×48 pixel and 2 channel (DAPI and brightfield) images. During training images were randomly rotated by 45-degrees and mirrored vertically or horizontally per iteration. In all CNN classifications, 48×18 pixel sub-images around each nuclei-center were generated. Cells closer than 25 pixels to the border of an image were excluded from all classifications.

### Drug response analysis & response prediction

Relative blast fractions (RBFs) were calculated as the fraction of cells of population X (identified by the CNN) after drug treatment divided by the average fraction of population X cells measured in control wells. Antibody-based treatments were normalized towards their respective isotype control whereas all other drugs were normalized towards DMSO. For calculation of *ex vivo* drug responses, all RBF values per sample were averaged over technical replicates, and subsequently zero-centered (1–RBF). Thus, a positive score represents a relative reduction of that population (on-target effect). If a drug eradicates all cells of this population without reducing other populations, the RBF score will be equal to 1. A RBF negative score indicates relative *ex vivo* chemoresistance of that population and killing all non-target cells, the score goes to negative infinity however values will be capped at -1. If a drug destroys both target and non-target cell populations at an equal proportion or has no effect on either population, the score becomes 0.

Mean maturation scores were calculated by averaging the RBF values of all present target cell populations. A cell population was defined as being present in a sample if it had a fraction higher than 0.005. Specifically, RBF values of the *H0, H1, H2, H3, Negs, All marker+, T-cell* and the *total cell number* reduction were taken into account. To punish the reduction of the healthy T-cell population, T-cell RBF were sign changed before averaging (hence reducing the T-cell population will negatively affect the score). The rationale behind this score is to screen for compounds which specifically kill all the malignant target cell populations, irrespective of their relative abundances while largely sparing healthy non-target cells, because even small malignant populations can negatively affect treatment outcome. A positive score favors drugs which relatively reduce all target populations whereas a negative score indicates incomplete target populations reduction.

### Statistical analysis and visualizations

Statistical analysis and visualization of data was performed using RStudio (v4.1.0) and Matlab R2021a. If not stated otherwise, all p-values were calculated based on wilcoxon rank sum test. The distribution of overall and event-free survival in the different subgroups was estimated using the Kaplan-Meier method and p-values for the survival analysis were calculated with the Gehan-Breslow-Wilcoxon Test.

## Data availability statement

CNN training and test datasets are available at: https://doi.org/10.3929/ethz-b-000680077. RNA-seq measurements used in the study are available at the GEO repository at: https://www.ncbi.nlm.nih.gov/geo/query/acc.cgi?acc=GSE272681. The *ex vivo* response data will be made available upon request. Additional data generated in this study are not publicly available due to information that could compromise patient privacy or consent but are available upon reasonable request from the corresponding author.

## Supplementary figure legends

**Figure S1.**
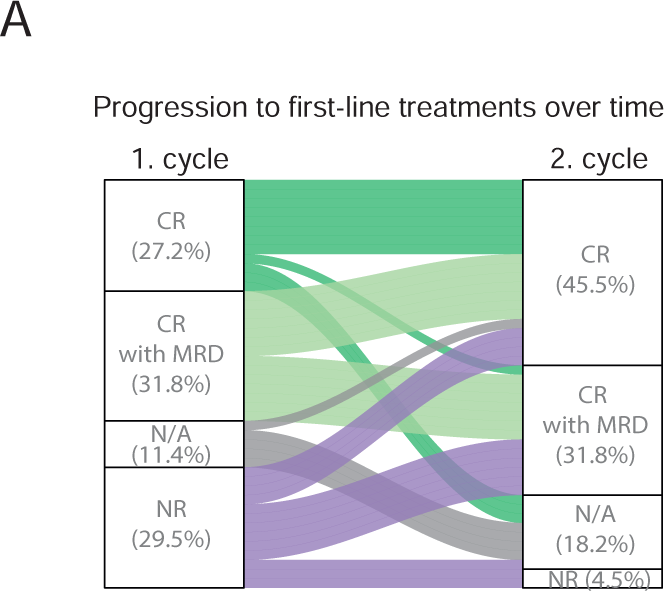
**A**, Progression to first line treatment responses over time. Flow diagram shows the clinical response to the first and the subsequent second induction chemotherapy for each patient (n=44). CR=complete remission. MRD=minimal residual disease. NR=non responder. N/A no response available.

**Figure S2.**
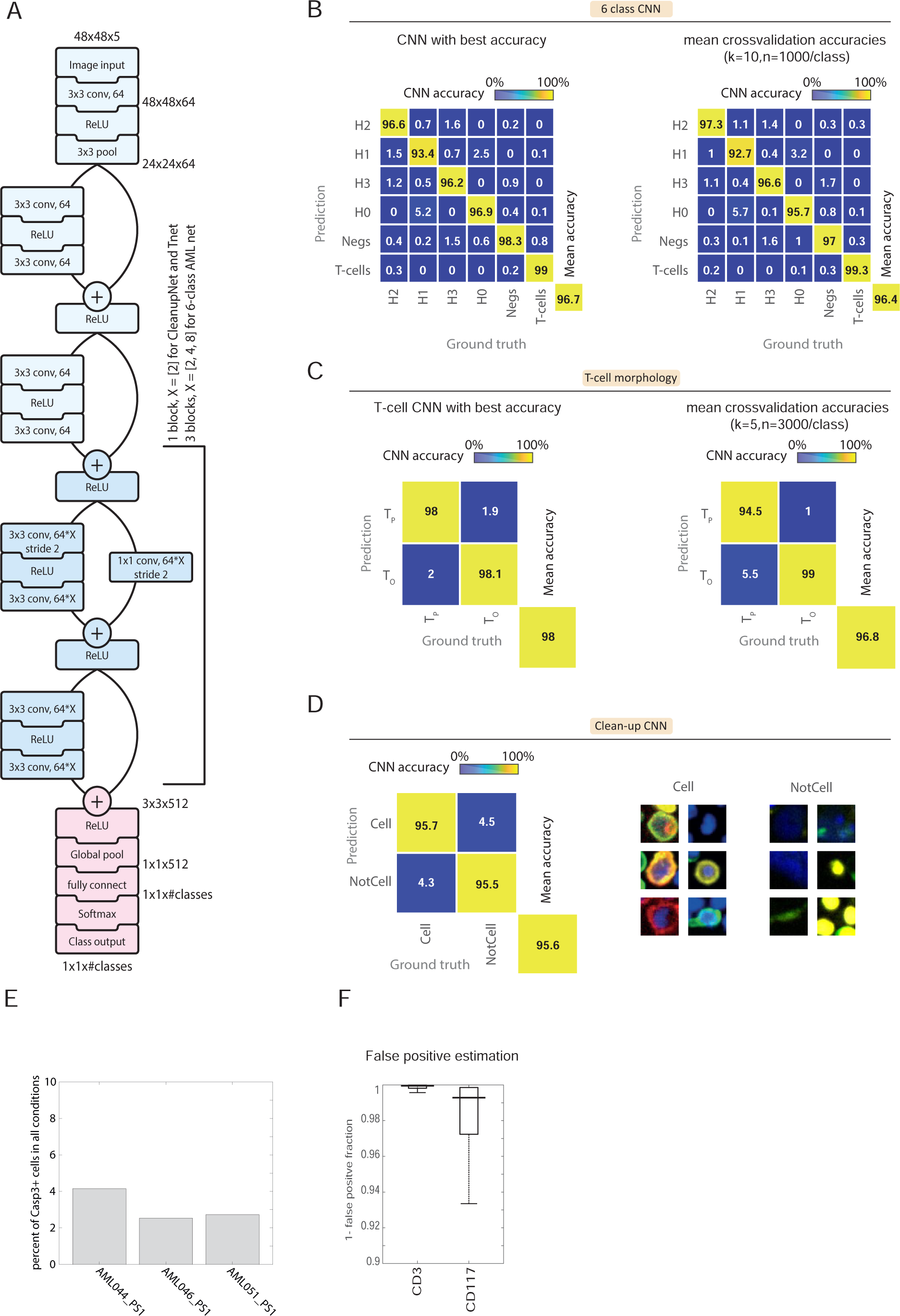
**A**, Schematic of the adapted ResNet architecture utilized in this study. **B,** Confusion matrix of the 6-class CNN performance across 1000 test cells per class. CNN was trained and tested 10 times with a random set of test cells (10% of dataset). Left shows the network with the best training & test performance which was used in this study. Right shows the average accuracies for all 10 runs. **C,** Confusion matrix of the T-cell morphology network across 3000 test cells per class. CNN was trained and tested 5 times. Left shows the network with the best training & test performance which was used in this study. Right shows the average accuracies for all 5 runs. **D,** Left: Confusion matrix of cleanup CNN. CNN was tested on 6000 cells per class (10% of data). Right: Representative example images of curated cells for cell and non-cellular object. The size of image crops is 48×48 pixels (14.4×14.4um). **E,** Percentage of apoptotic cells after cleanup. Bar Graph shows the average percentage of Casp3+ positive cells after cleanup by the ‘Cleanup CNN’ across a full drug plate for three individual patients. **F,** False-positive classification of the 6-class CNN on non-multiplexed test cells. Each datapoint in the boxplot diagram represents one sample (n=174). False positive score is calculated as 1-(fraction of misclassified cells per control per sample).

**Figure S3.**
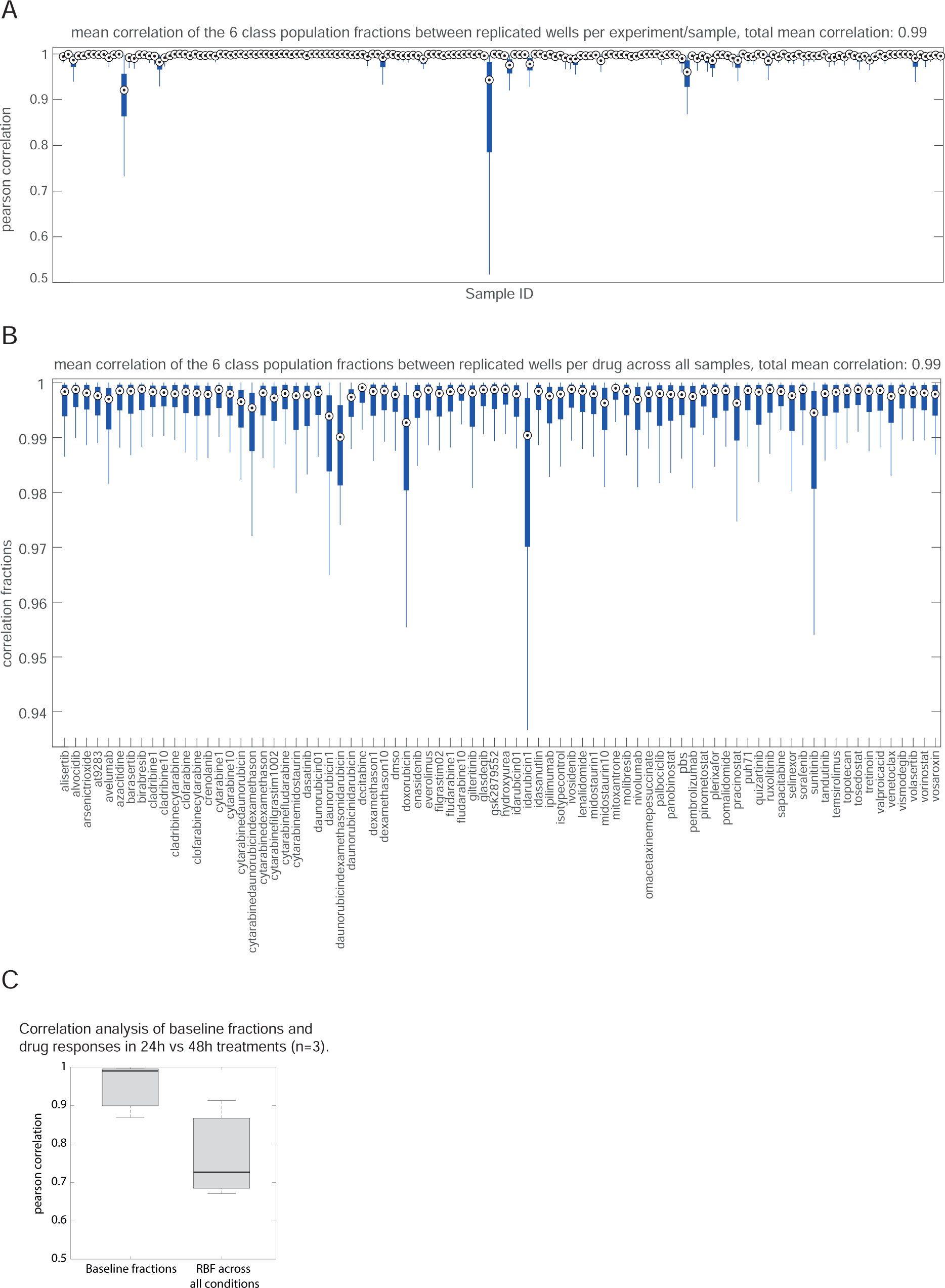
**A**, Technical reproducibility per sample. The pairwise correlation of mean population fractions across all technical replicates per sample per control is calculated. All calculated correlations are depicted as boxplots. **B,** Technical reproducibility per drug. The average pairwise correlation of mean population fractions per individual drug in each sample is calculated. Boxplots depict correlations for all 174 samples. **C,** Correlation of 24h treatment vs 48h treatment across all population drug responses for three individual patients at diagnosis. For all boxplots the black dot indicates the median. Bottom and top edges of the black box indicate the 25th and 75th percentiles, respectively. The whiskers extend to the most extreme data points not considered outliers whereas the outliers are plotted in red.

**Figure S4.**
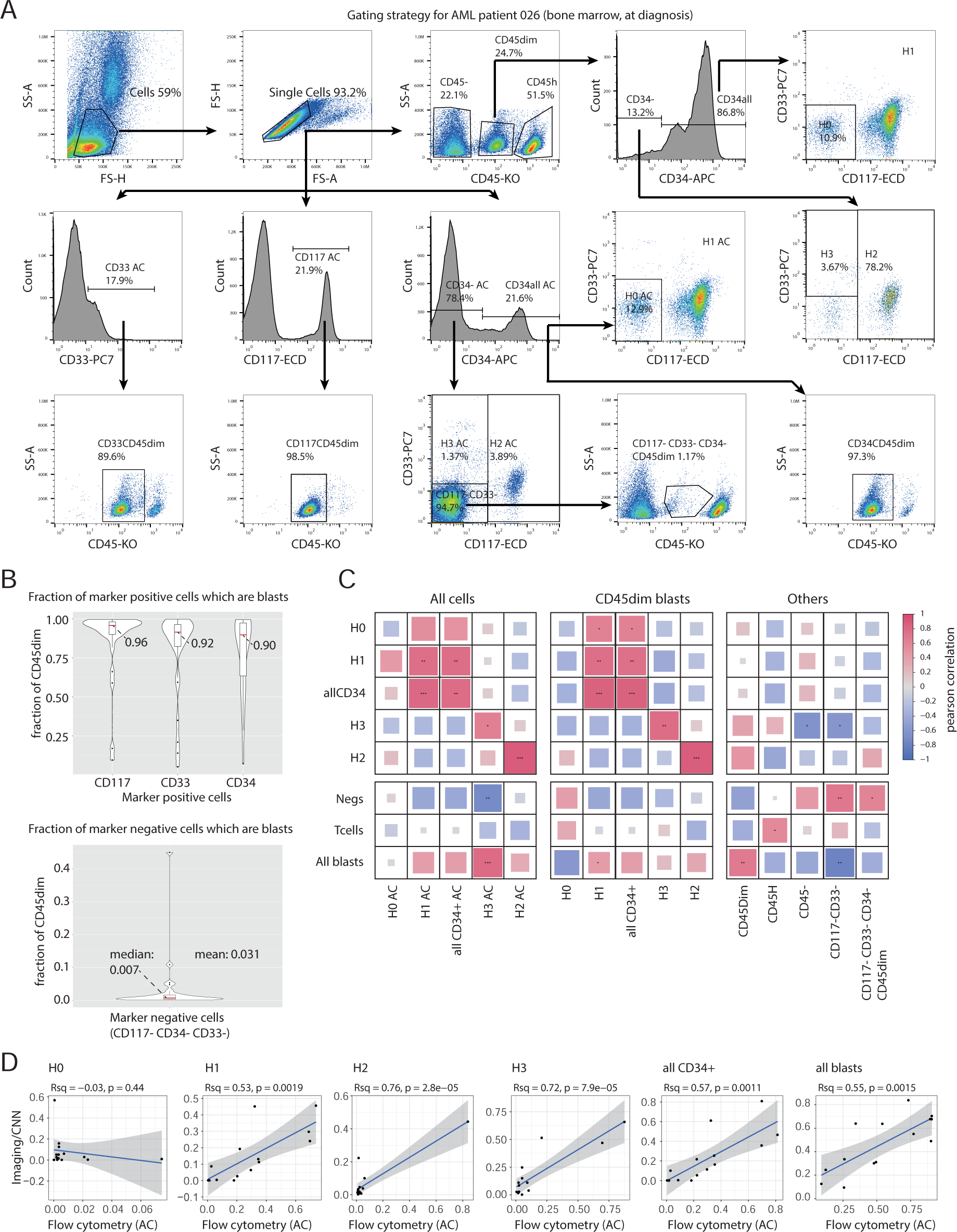
**A**, Gating strategy for matching population definitions by flow cytometry. 27 samples at diagnosis with matched CD34, CD117, and CD33 staining were analyzed. **B,** Upper: Violin plots showing fraction of CD117, CD33 and CD34 marker positive cells which fall within the CD45dim (blast) gate. Lower: Violin plot showing the fraction of CD117-CD33-CD34-triple negative cells which fall within the CD45dim (blast) gate. Measurements were conducted by flow cytometry at diagnosis. **C,** Pearson correlation of populations abundances measured at diagnosis measured by either flow cytometry or Imaging/Pharmacoscopy. **D,** Scatter plot correlating population abundances measured by either flow cytometry or imaging. The blue line represents a linear model fit, with p-value and Rsq value from the linear regression indicated. P Value asterisks are defined as following: *p<0.05, **p<0.01, ***p<10^-3^.

**Figure S5.**
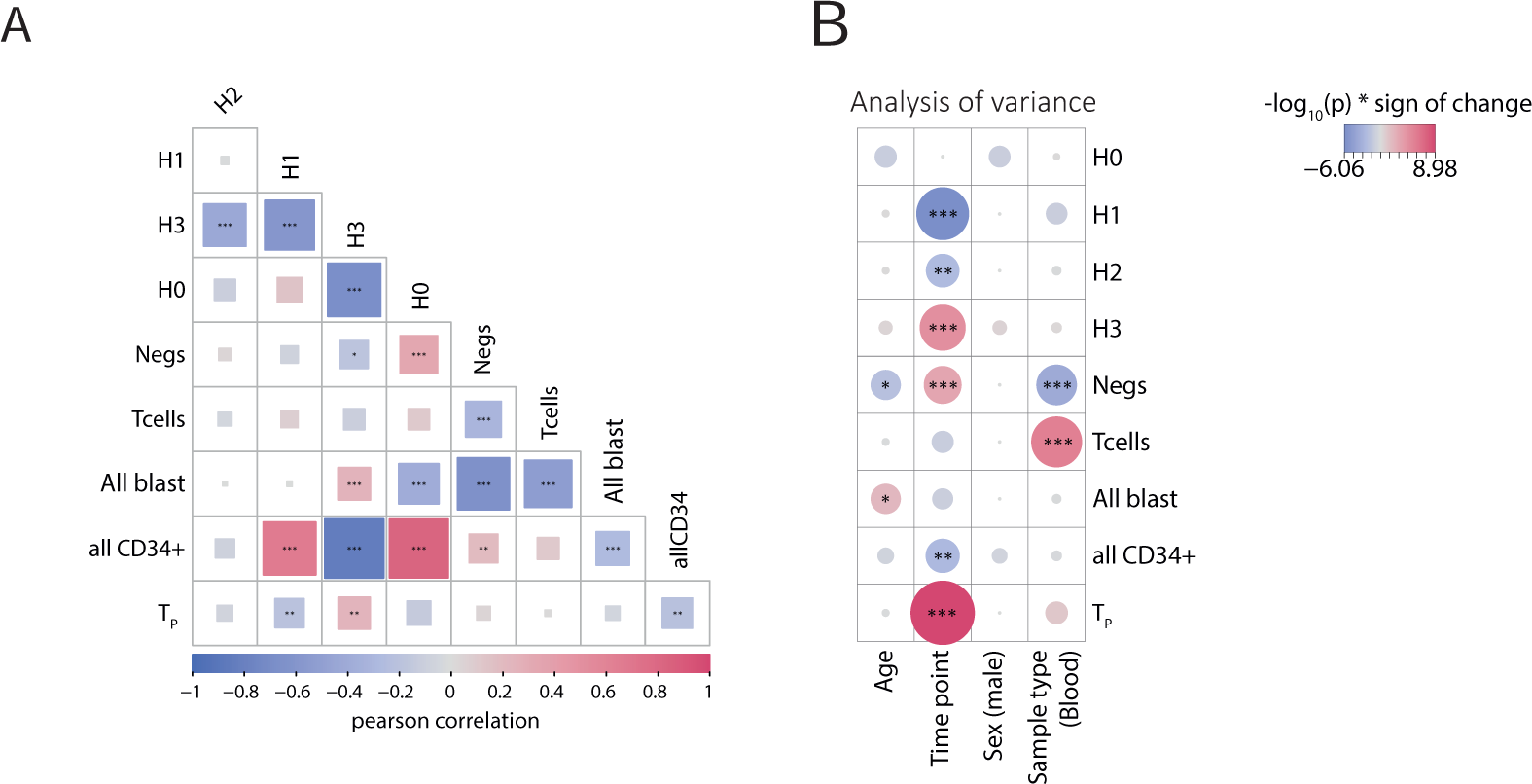
**A**, Correlation of population abundances at baseline. Mean population fractions were pearson correlated across all samples (n=174). All shown fractions were obtained from DMSO treated control wells. **B,** Heatmap overview of individual influences on population composition. Significance of influence was calculated by analysis of variance based on population abundances shown in Figure 2A. P-values were log_10_ transformed and multiplied by the sign of change. For all calculations H0, H1, H2, H3, all CD34+ population fractions were normalized towards the absolute AML abundance. P-value asterisks are defined as following: *p<0.05, **p<0.01, ***p<10^-3^.

**Figure S6.**
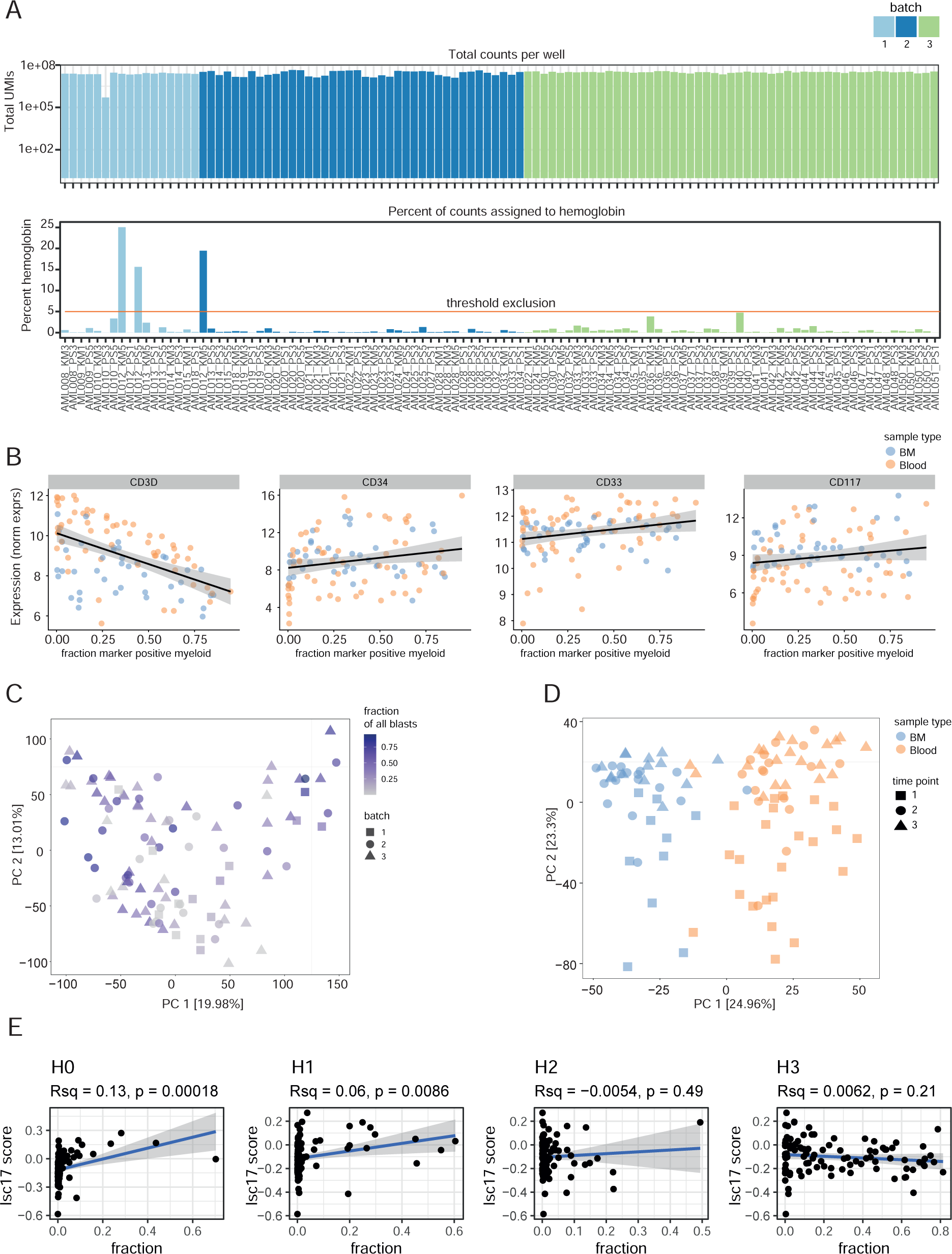
**A**, Top: Bar graphs indicated the sum of transcript counts after DESeq2 normalization ^46^. Color indicates the sequencing batch. n=108 samples. Bottom: Bar graphs indicated the percent of transcript count assigned to hemoglobin. Samples above a threshold of 5% of hemoglobin (and with a lower gene count than 14000 or a lower minimal count than = 10^6^) were excluded from analysis (total of 4 samples). **B,** Scatter plot of selected normalized transcript expression values per sample against the fraction of all marker positive AML cells. Color by tissue origin of sample. **C,** Principal component analysis of normalized transcript values of all samples colored by the fraction of marker positive AML cells. Shapes indicate batch. **D,** Principal component analysis of normalized and corrected transcript values of all samples colored by tissue origin of sample. Transcript values were corrected for the fraction of all marker positive AML cells. Shapes indicate time point. **E,** Scatter plot of the LSC17 stemness score calculated per sample against AML population fraction of each sample. Blue line indicates the fit of a linear model. P-value and Rsq of the linear regression are shown. All marker positive AML cells are defined to be either CD33, CD34 or CD117 positive. BM = bone marrow.

**Figure S7.**
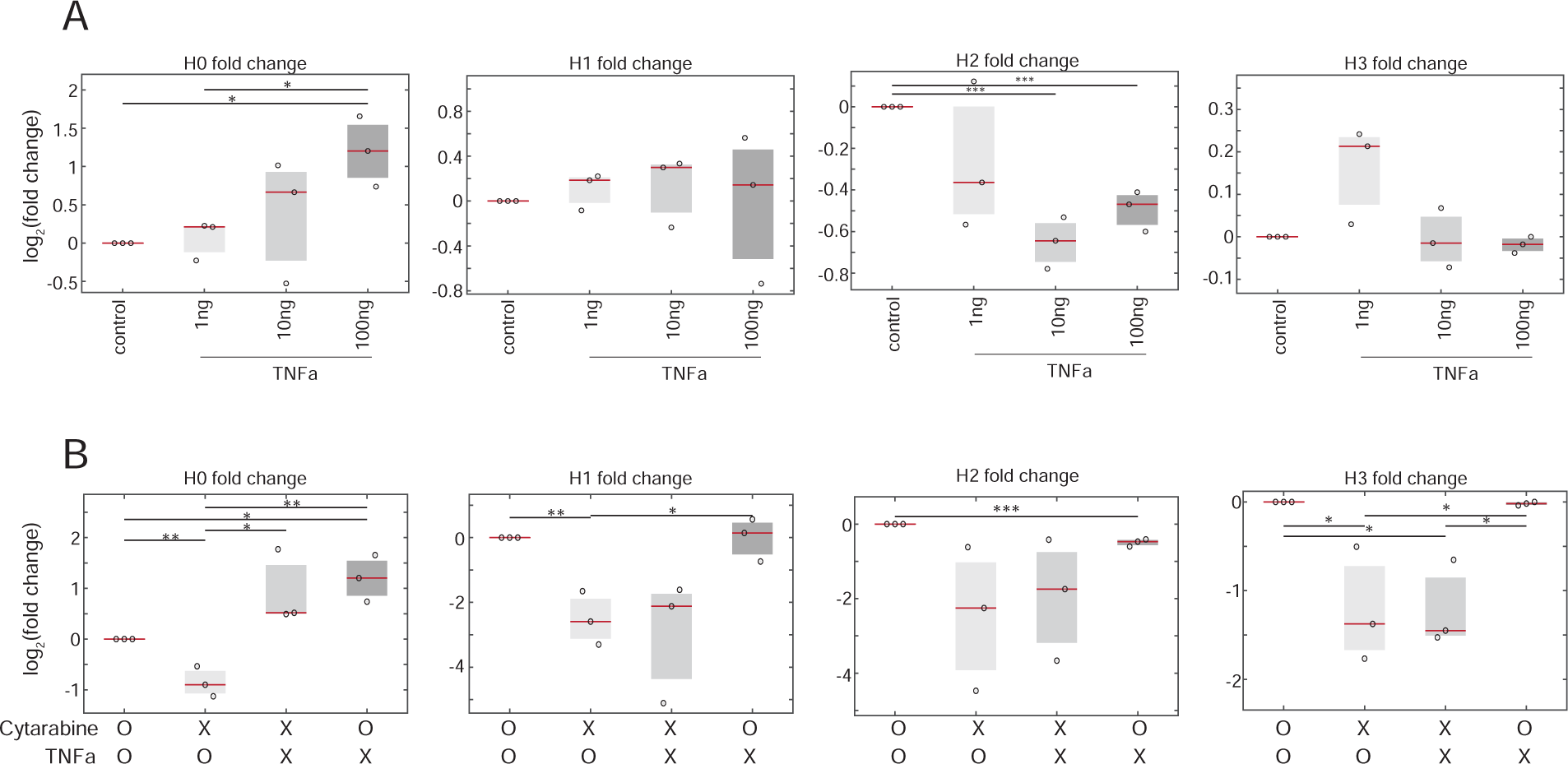
Induction and suppression of the H0 phenotype with TNF-α and cytarabine treatment. Plotted are the log_2_ fold changes of the H0 fraction across three individual AML patients at diagnosis compared to control treatments. **A,** Increasing concentrations of TNF-α treatment for 48h. Values were normalized against a PBS control**. B,** Combinations of cytarabine (10um) and TNF-α (100ng) against matching controls. P-values were calculated based on a two-tailed Student’s *t*-test.

**Figure S8.**
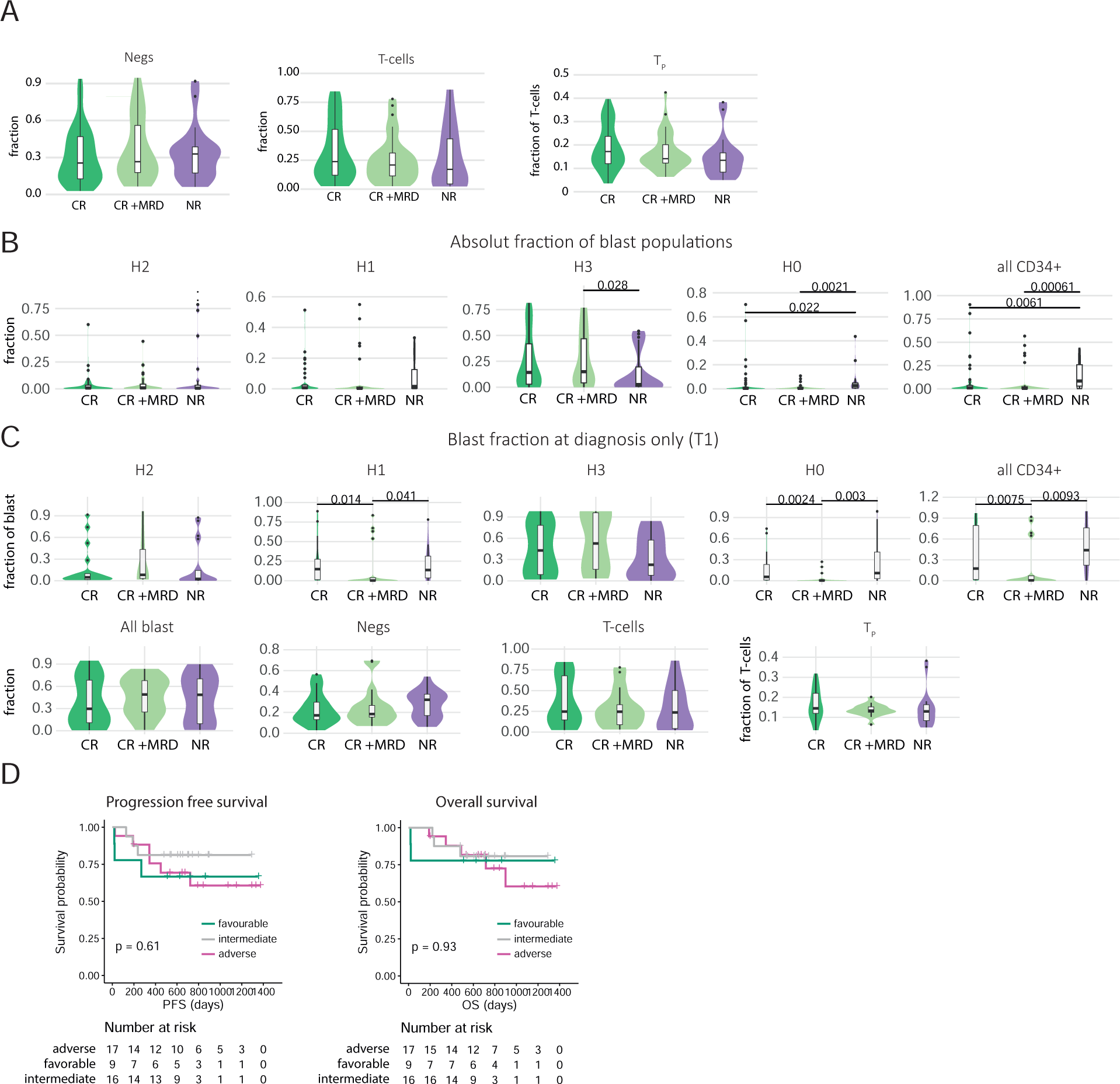
**A**, Population abundance in relation to treatment outcome. Violin plot displaying the distribution of different population fractions measured before clinical treatment response (first or second round). **B,** Violin plot displaying the distribution of different absolute population fractions measured before clinical treatment response (first or second round). **C,** Violin plot displaying the distribution of different population fractions measured at diagnosis. Population fractions were normalized towards the absolute AML abundance. **D,** ELN Risk score stratification (n=42 patients) plotted as Kaplan-Meier survival curves. Samples with no response were excluded from analysis. CR=complete remission. MRD=minimal residual disease. NR=non responder.

**Figure S9.**
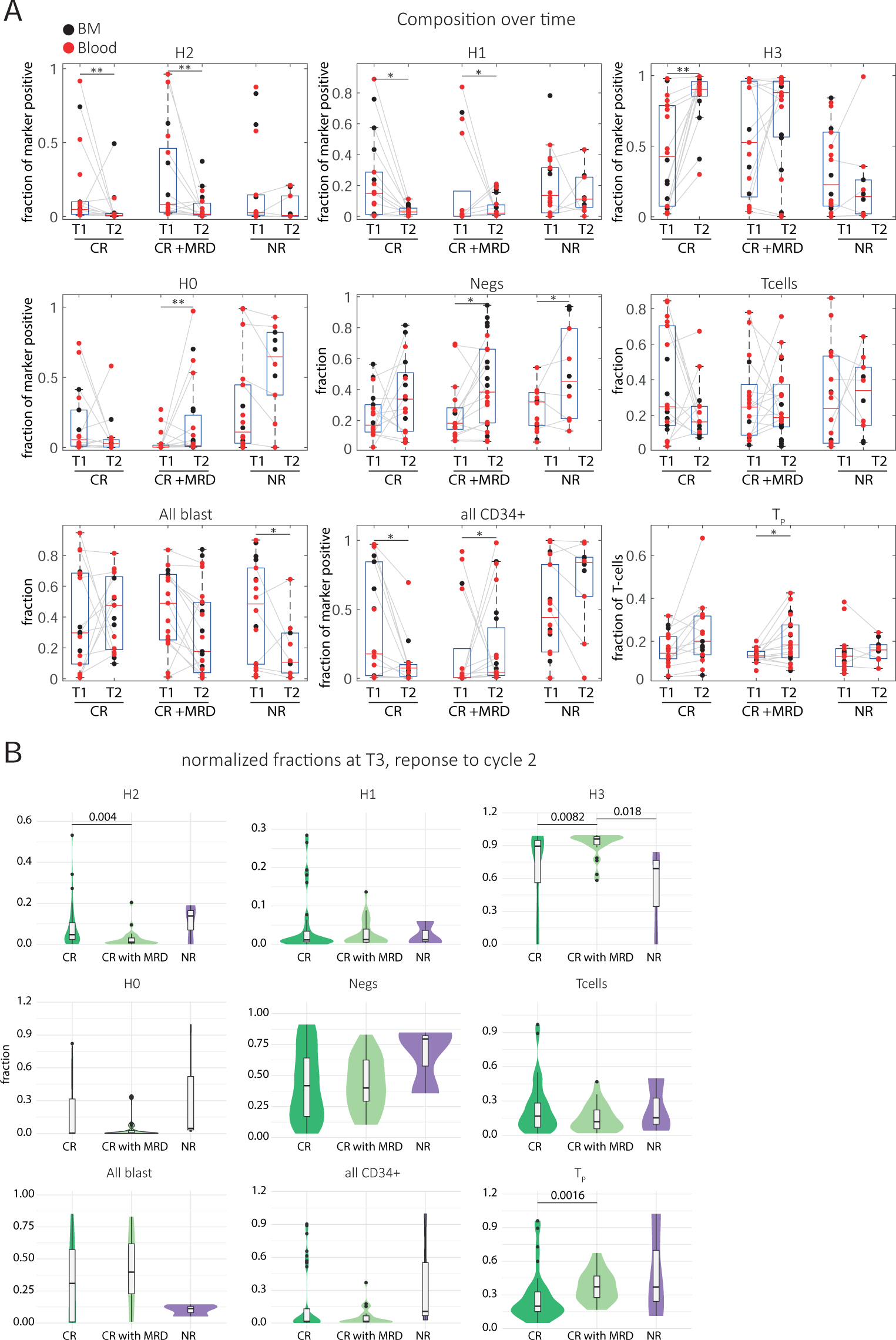
**A**, Population composition over time. Box plot displaying the fraction of different population abundances measured before and after the first round of treatment stratified by patient response. Grey lines connect matching samples from the same patient and tissue. AML populations were normalized towards the absolute AML abundance. **B,** Violin plot displaying the distribution of different population fractions measured at timepoint 3 (after second cycle). CR=complete remission. MRD=minimal residual disease. NR=non responder. *p<0.05, **p<0.01, ***p<10^-3^.

**Figure S10.**
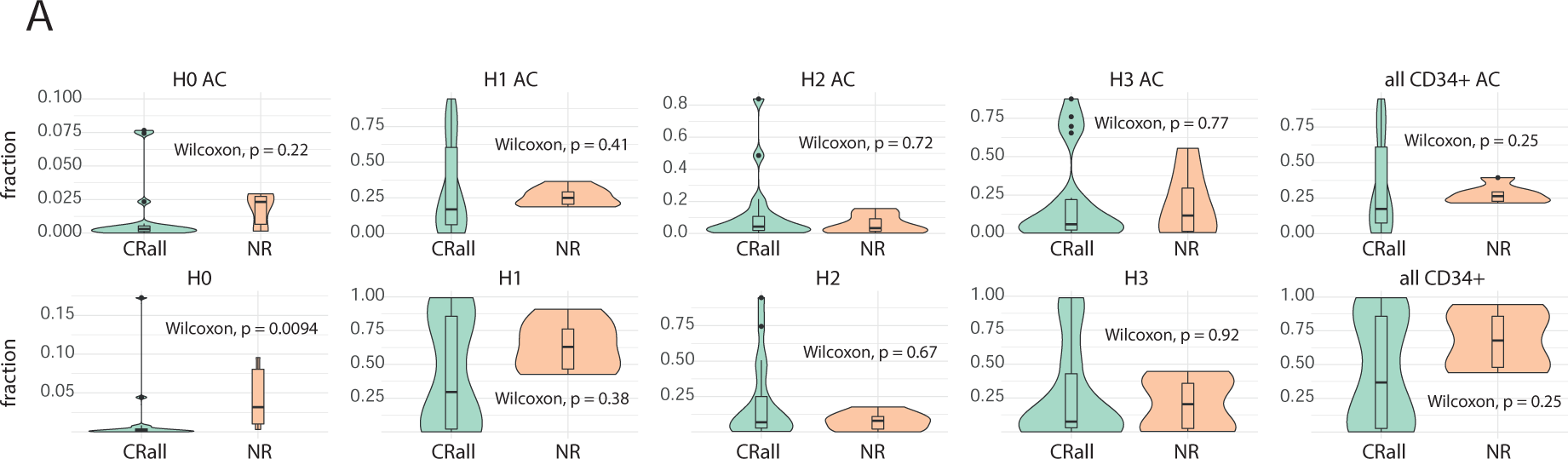
**A**, Population composition at diagnosis measured by flow cytometry. 27 samples at diagnosis with matched CD34, CD117, and CD33 staining were analyzed. Violin plot displaying the distribution of different AML population abundances. AML populations were normalized towards the absolute AML abundance. CR=complete remission. MRD=minimal residual disease. NR=non responder.

**Figure S11.**
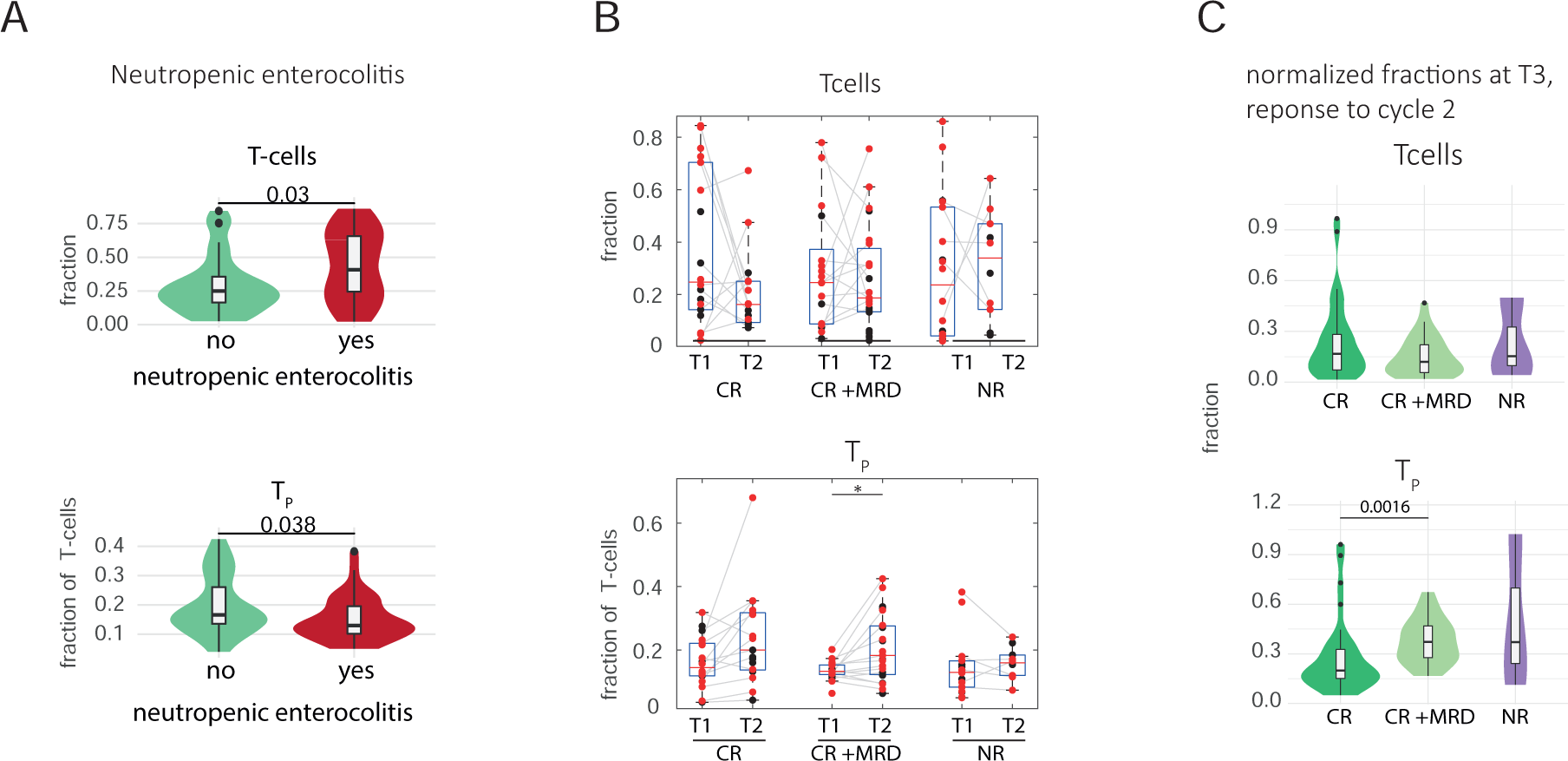
**A**, T-cell population abundance stratified by the emergence of neutropenic enterocolitis. Violin plot displaying the distribution of the absolute T-cell fraction measured at T1 & T3.**B,** T-cell population composition over time. Box plot displaying the fraction of different population abundances measured before and after the first round of treatment stratified by patient response. Grey lines connect matching samples from the same patient and tissue. **C,** Violin plot displaying the distribution of T-cell population fractions measured at timepoint 3 (after second cycle). CR=complete remission. MRD=minimal residual disease. NR=non responder. T_P_ were normalized towards the absolute T-cell abundance.

**Figure S12.**
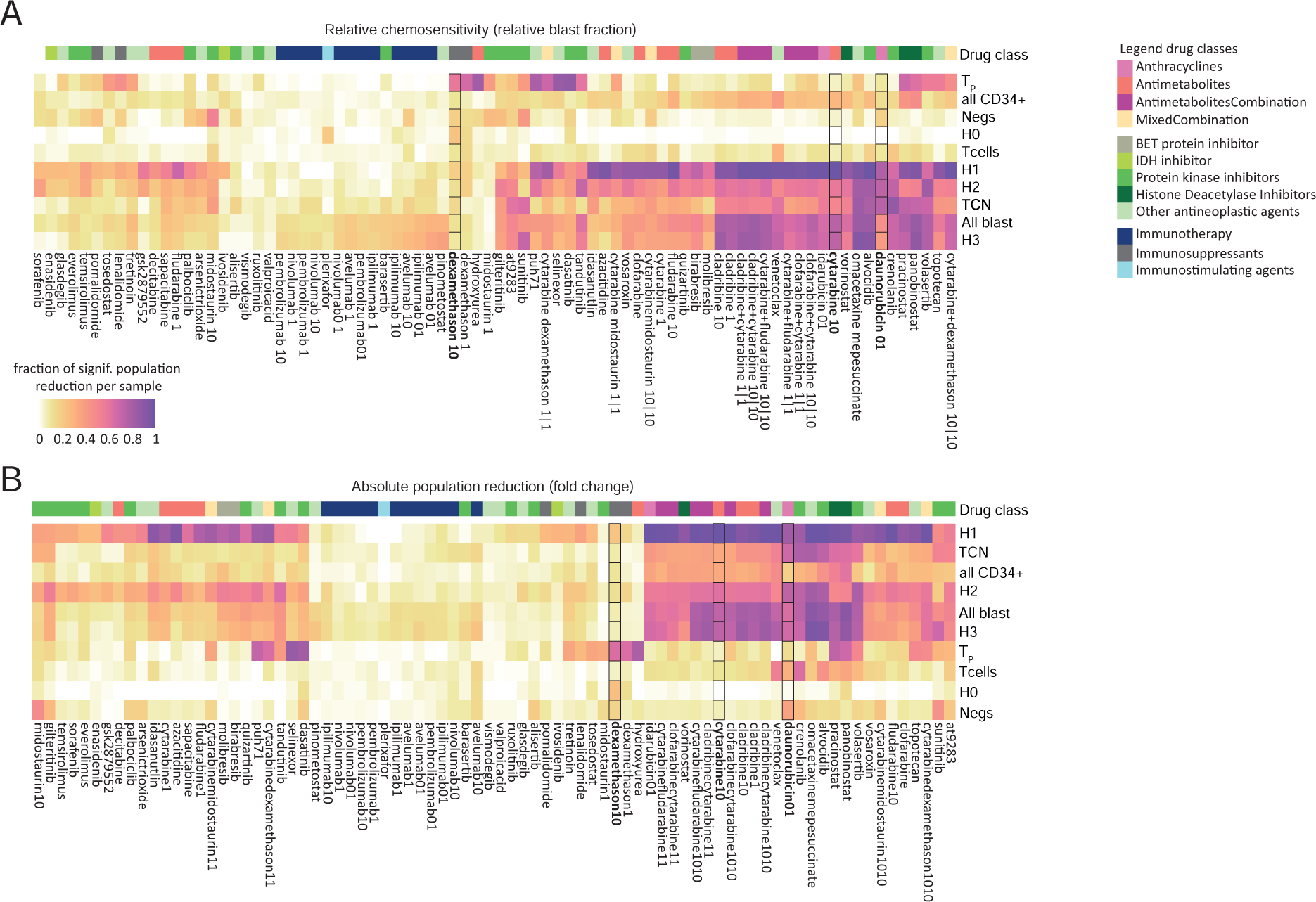
Drug response heterogeneity in relation to AML maturation stage. Heatmaps display the fraction of samples with a significant reduction in the target population in a given treatment. **A,** Fraction of significant relative abundance changes within the population (1-RBF) **B,** Fraction of significant absolute abundance changes within the population (fold change). N=80 treatments.

**Figure S13.**
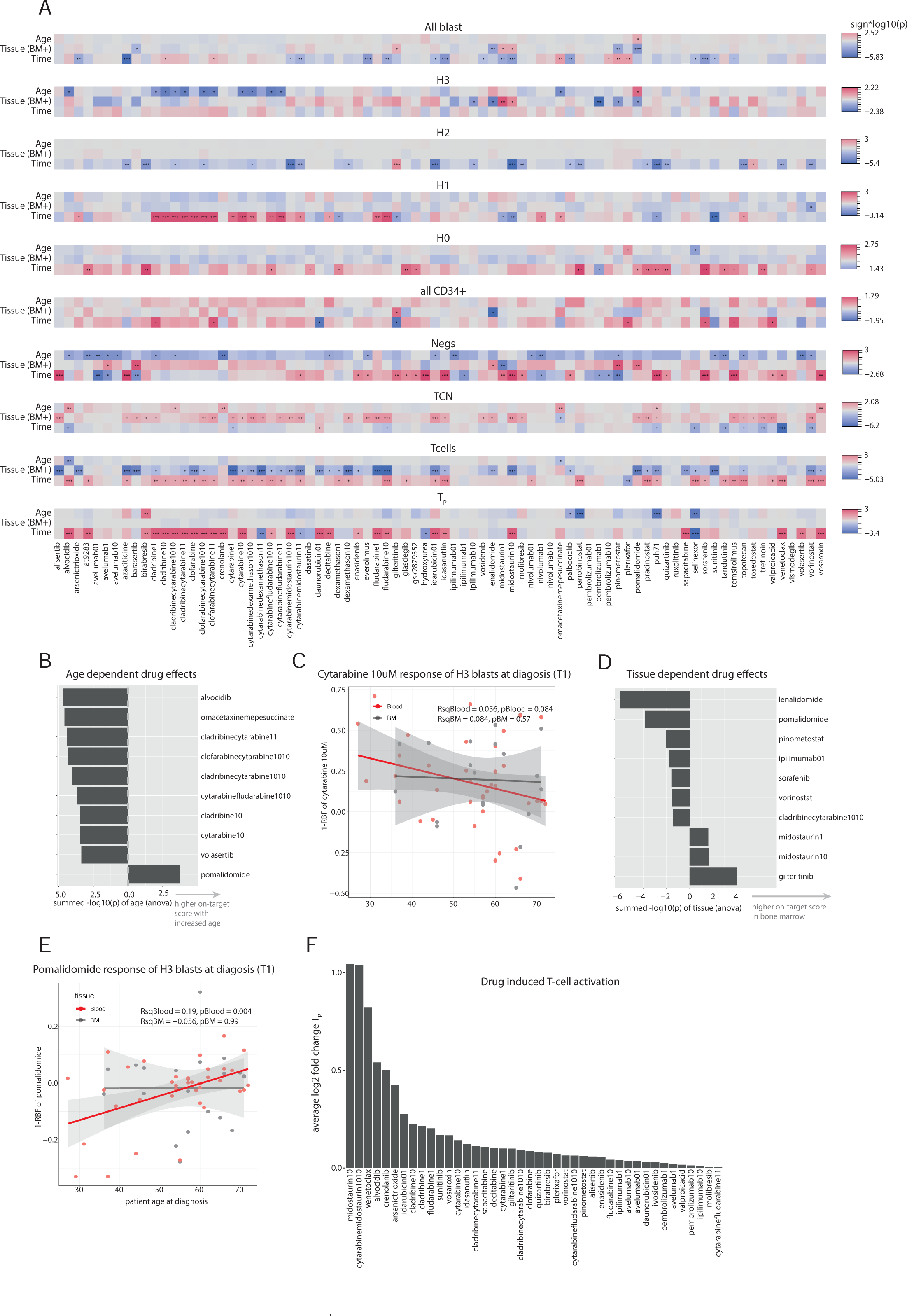
**A**, Heatmap overview of individual influences on drug responses stratified by population. Significance of influence was calculated by analysis of variance (ANOVA). P-values were log_10_ transformed and multiplied by the sign of change. *p<0.05, **p<0.01, ***p<10^-3^. **B,** Identification of the Top 10 drugs exhibiting the most pronounced age-dependent responses. Age-dependent response is quantified as the summed p-values (directed -log10(p)) of age scores calculated in A, across various cell populations, including H0-H3 stages, negative controls (Negs), all blast cells, and all CD34+ progenitor cells. Positive values indicate responses with higher on-target scores with increased age. **C,** Scatter plot of the H3 population response to cytarabine 10uM in relation to patient age stratified by tissue. Only samples at diagnosis were considered. Line indicates the fit of a linear model. p value and Rsq of the linear regression are shown per tissue. **D,** Identification of the Top 10 drugs exhibiting the most pronounced tissue-dependent responses. Tissue-dependent response is quantified as the summed p-values (directed -log10(p)) of scores calculated in A, across various cell populations, including H0-H3 stages, negative controls (Negs), all blast cells, and all CD34+ progenitor cells. Positive values indicate responses with higher on-target scores in bone marrow samples. **E,** Scatter plot of the H3 population response to pomalidomide (10uM) in relation to patient age stratified by tissue. Only samples at diagnosis were considered. Line indicates the fit of a linear model. P-value and Rsq of the linear regression are shown per tissue. **F,** Quantification of drug-induced T-cell activation. The ranked bar plot presents the average log2 fold changes of the T_P_ population calculated across all samples per drug. Only positive scores are shown.

**Figure S14.**
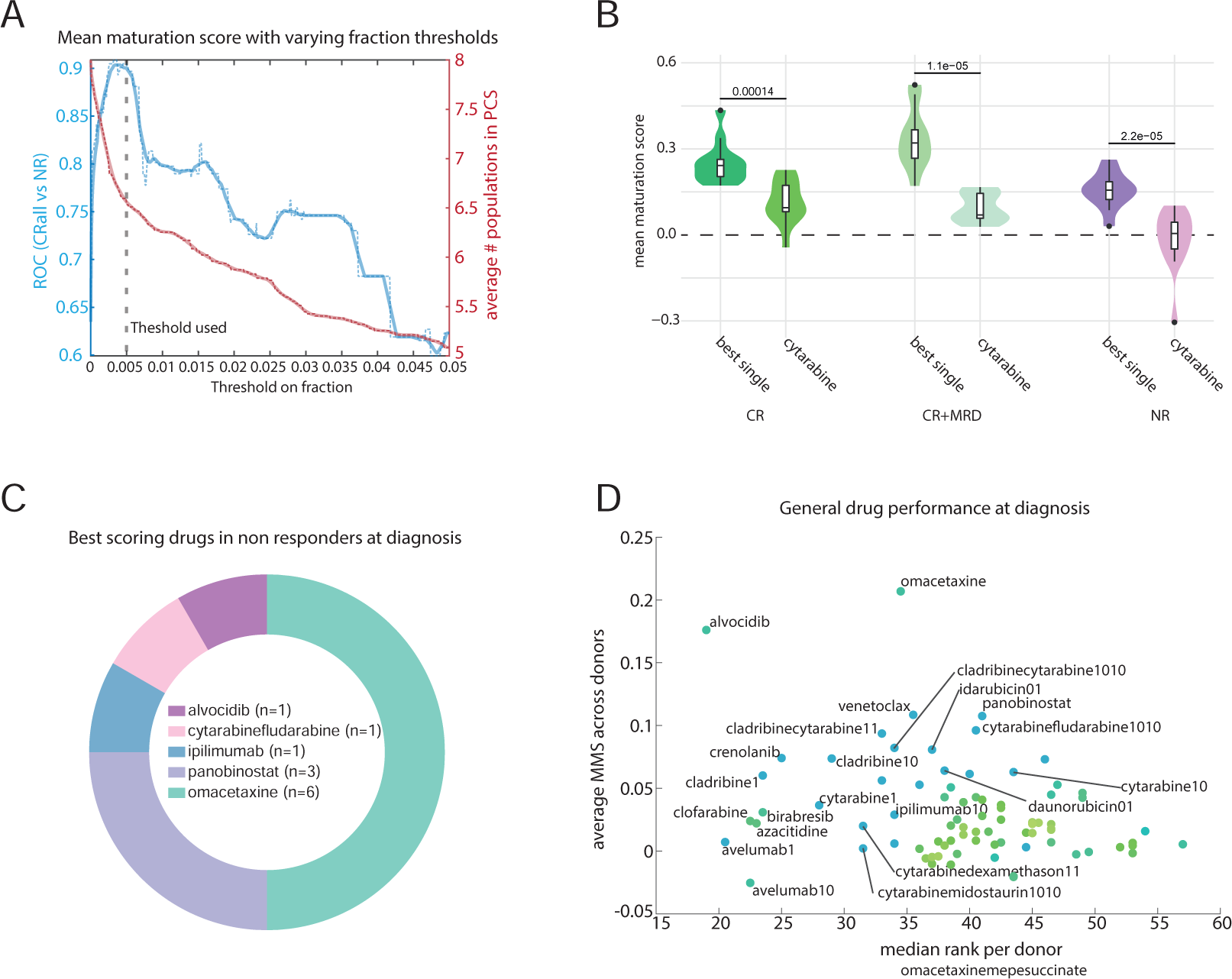
**A**, Stratification of responders and non-responders to first line treatment by mean maturation scores with varying factions thresholds. Mean maturation scores were calculated by averaging the RBF values of all present target cell populations. A cell population was defined as being present in a sample if it had a fraction higher than the specified threshold (x-axis). Negs were including the neg as a target population (see methods). Left y axis (blue): Area under curve of the ROC curve for the optimal MMS cutoff at a given fraction threshold stratifying responders from non-responders. Right y axis (red) average number of populations included in the MMS across all samples with a given fraction threshold. **B,** Stratification of MMS Scores at diagnosis by the ex vivo response to cytarabine and the single treatment with the highest MMS score per sample. Violin plot illustrates the distribution of MMS scores measured at the point of diagnosis, categorized according to the patients’ clinical response to first line treatment. Measurements were taken before a patient was treated. Patients without clinical response annotation were excluded from this analysis. CR=complete remission. MRD=minimal residual disease. NR=non responder. **C,** Best scoring treatments in non-responding patients at diagnosis. **D,** General drug performance at diagnosis. Scatter plot showing the median drug rank across patients vs the average MMS score. Drugs were ranked by their average MMS within each patient (with 1 being the rank of best performing drug in a given sample). If multiple tissues (blood and bone marrow) were measured per patient, the average MMS score across both samples was calculated first.

**Figure S15.**
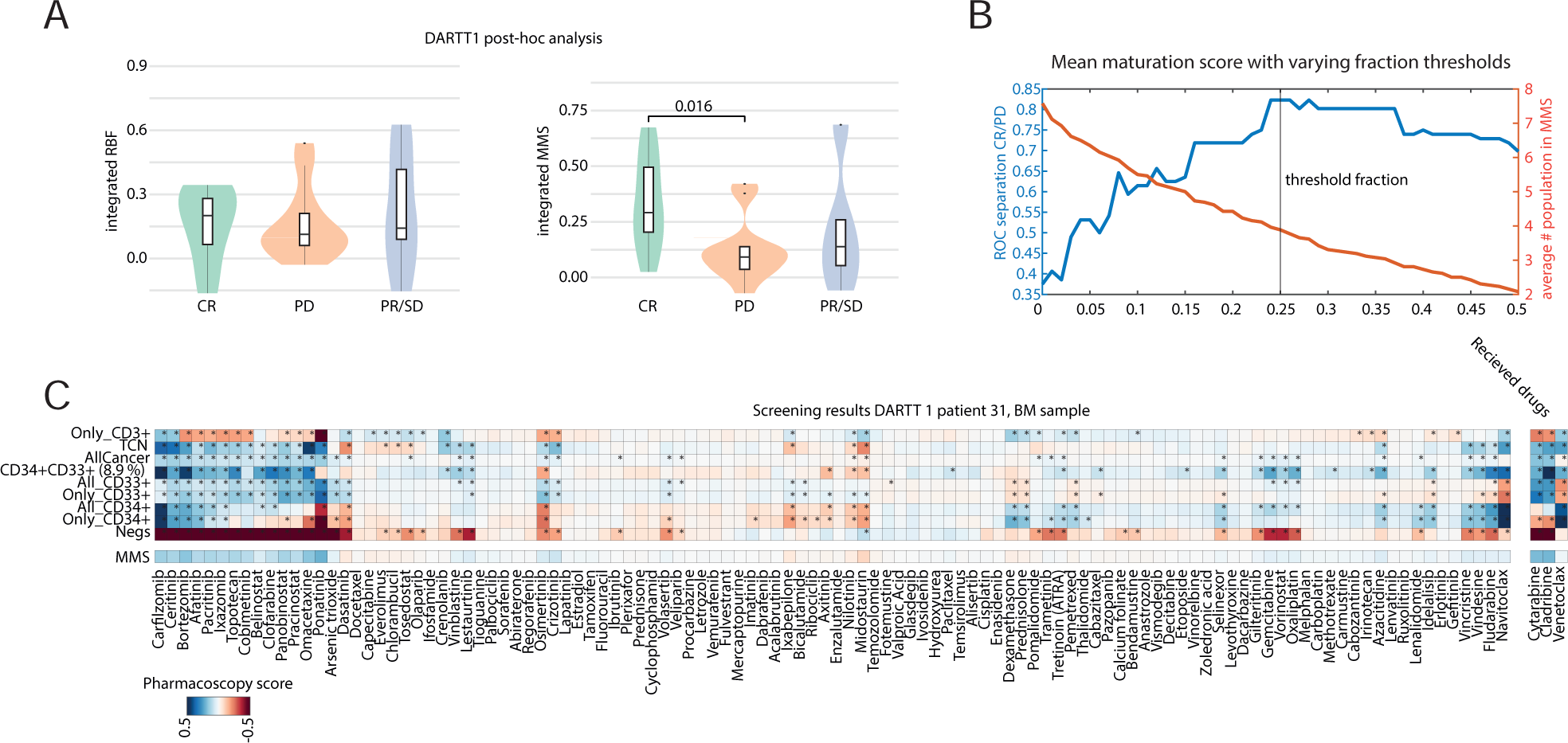
**A**, DARTT-1 post-hoc analysis. Violin plot shows the integrated ex vivo RBF and MMS scores stratified by the clinical treatment response. Integrated MMS score was calculated by summing all MMS scores for every individual chemotherapy a patient was treated with. Measurements were taken before a patient was treated. CR=complete remission. PD=progressive disease. PR: Partial response. SD: Stable disease. **B,** Stratification of patient samples achieved a complete remission (CR) or faced a progressive disease (PD) treatment by MMS with varying fractions thresholds. Mean maturation scores were calculated by averaging the RBF values of all present target cell populations. A cell population was defined as being present in a sample if it had a fraction higher than the specified threshold (x-axis). Negs were including the neg as a target population (see methods). Left y axis (blue): Area under curve of the ROC curve for the optimal MMS cutoff at a given fraction threshold stratifying responders from non-responders. Right y axis (red) average number of populations included in the MMS across all samples with a given fraction threshold. **C,** Example screening results of DART 1 patient 31. Heatmap is ranked by MMS score.

## Contributions

Conceptualization: Y.S., M.S., S.S., M.G.M.., A.M.S.M., B.S. Methodology: Y.S., Y.F., T.M.B. Software: Y.S., R.W. Validation: Y.S., Y.F. Formal analysis: Y.S., Y.F., R.W., B.D.H. Investigation: Y.S., Y.F. Resources: T.M.B., M.R., A.K.K., T.P., M.S. Data Curation: Y.S., Y.F., T.M.B., B.D.H. Writing - Original Draft: Y.S., Y.F., B.S. Writing - Review & Editing: Y.S., Y.F., T.M.B., R.W., B.D.H., M.G.M., S.S., A.M.S.M., B.S. Visualization: Y.S. Supervision: Y.S., S.S., M.S., M.G.M., B.S. Project administration: Y.S., A.M.S.M., M.G.M., S.S., B.S. Funding acquisition: S.S., M.G.M., A.M.S.M., M.S., B.S.

## Acknowledgments

We thank the anonymous patients for their contributions to this study. We also acknowledge the support of Tim Heinemann, Melanie Clerc, Luca Truscello, and all members of the Snijder laboratory for their discussions and additional experimental assistance.

